# Improving Precision and Power in Randomized Trials for COVID-19 Treatments Using Covariate Adjustment, for Binary, Ordinal, and Time-to-Event Outcomes

**DOI:** 10.1101/2020.04.19.20069922

**Authors:** David Benkeser, Iván Díaz, Alex Luedtke, Jodi Segal, Daniel Scharfstein, Michael Rosenblum

## Abstract

Time is of the essence in evaluating potential drugs and biologics for the treatment and prevention of COVID-19. There are currently over 400 clinical trials (phase 2 and 3) of treatments for COVID-19 registered on clinicaltrials.gov. Covariate adjustment is a statistical analysis method with potential to improve precision and reduce the required sample size for a substantial number of these trials. Though covariate adjustment is recommended by the U.S. Food and Drug Administration and the European Medicines Agency, it is underutilized, especially for the types of outcomes (binary, ordinal and time-to-event) that are common in COVID-19 trials. To demonstrate the potential value added by covariate adjustment in this context, we simulated two-arm, randomized trials comparing a hypothetical COVID-19 treatment versus standard of care, where the primary outcome is binary, ordinal, or time-to-event. Our simulated distributions are derived from two sources: longitudinal data on over 500 patients hospitalized at Weill Cornell Medicine New York Presbyterian Hospital, and a Centers for Disease Control and Prevention (CDC) preliminary description of 2449 cases. We found substantial precision gains from using covariate adjustment-equivalent to 9-21% reductions in the required sample size to achieve a desired power-for a variety of estimands (targets of inference) when the trial sample size was at least 200. We provide an R package and practical recommendations for implementing covariate adjustment. The estimators that we consider are robust to model misspecification.

## 1. Introduction

This paper builds on our report (Benkeser et al., 2020) written in response to a request by the U.S. Food and Drug Administration (FDA) for statistical analysis recommendations for COVID-19 treatment trials. We aim to help inform the choice of estimand (i.e., target of inference) and analysis method to be used in future and ongoing COVID-19 treatment trials. To this end, we describe treatment effect estimands whose interpretability does not rely on correct specification of models and that may be of particular interest in COVID-19 treatment trials involving binary, ordinal, or time-to-event outcomes. For binary outcomes, we consider the risk difference, relative risk, and odds ratio. For ordinal outcomes, we consider the difference in means, the Mann-Whitney (rank-based) estimand, and the average of the cumulative log odds ratios over levels of the outcome. For time-to-event outcomes, we consider the difference in restricted mean survival times, the difference in survival probabilities, and the ratio of survival probabilities. For each estimand, we present covariate adjusted estimators of these quantities that leverage information in baseline variables to improve precision and reduce the required sample size to achieve a desired power. In the case of ordinal outcomes, we propose novel covariate adjusted estimators.

To evaluate the performance of covariate adjusted estimators, we simulated two-arm, randomized trials comparing a hypothetical COVID-19 treatment versus standard of care, where the primary outcome is binary, ordinal, or time-to-event. Our simulated distributions are derived from two sources: longitudinal data on over 500 patients hospitalized at Weill Cornell Medicine New York Presbyterian Hospital prior to March 28, 2020, and a preliminary description of 2449 cases reported to the CDC from February 12 to March 16, 2020. We focused on hospitalized, COVID-19 positive patients and considered the following outcomes: intensive care unit (ICU) admission, intubation with ventilation, and death. We conducted simulations using all three estimands when the outcome is ordinal, but only evaluated the risk difference when the outcome is binary and the restricted mean survival time and risk difference when the outcome is time-to-event.

In our simulations of trials with at least 200 participants, the precision gains due to covariate adjustment were often substantial, equivalent to requiring 9-21% fewer participants to achieve the same power as a trial that uses an unadjusted estimator. In our simulated trials with 100 participants, covariate adjustment still offered improvements in precision, equivalent to requiring 4-18% fewer participants to achieve the same power as a trial that uses an unadjusted estimator. From these simulations, we conclude that covariate adjustment is a low-risk, high-reward approach to streamlining COVID-19 treatment trials.

After our aforementioned report (which contains the simulation results described above for ordinal and time-to-event outcomes), the FDA released a guidance for industry on COVID-19 treatment and prevention trials (FDA, 2020). The guidance contains the following statement, which is similar to our key recommendation regarding covariate adjustment: “To improve the precision of treatment effect estimation and inference, sponsors should consider adjusting for prespecified prognostic baseline covariates (e.g., age, baseline severity, comorbidities) in the primary efficacy analysis and should propose methods of covariate adjustment.”

There is already an extensive literature on the statistical theory and practice of covariate adjustment (e.g., Zhang et al. (2008); Jiang et al. (2019); Austin et al. (2010). However, covariate adjustment is underutilized, particularly for trials with a binary, ordinal, or time-to-event outcome. Since many COVID-19 treatment trials focus on exactly these types of outcomes, our goal with this work is to demonstrate, using real data, the potential benefits of covariate adjustment in these contexts.

The remainder is organized as follows. A background on covariate adjustment in randomized trials is provided in Section 2. Section 3 describes estimands and estimation strategies when the outcome is binary, ordinal, or a time-to-event. Section 4 describes the methods underlying the simulation study and Section 5 presents the simulation study results. Section 6 presents our recommendations for the estimands and primary efficacy analysis methods for future COVID-19 treatment trials. A brief discussion is given in Section 7.

## 2. Background on Covariate Adjustment in Randomized Trials

The ICH E9 Guidance on Statistical Methods for Analyzing Clinical Trials (FDA and EMA, 1998) states that “Pretrial deliberations should identify those covariates and factors expected to have an important influence on the primary variable(s), and should consider how to account for these in the analysis to improve precision and to compensate for any lack of balance between treatment groups.” The term “covariates” refers to baseline variables. Adjusting for pre-specified, prognostic baseline variables (i.e., variables that are correlated with the outcome) is called covariate adjustment.

Though there appears to be a general agreement among regulators (EMA, 2015; FDA, 2019) that when the outcome is continuous, analysis of covariance (ANCOVA) may be used to appropriately adjust for baseline variables, there is a dearth of specific guidance for ordinal and time-to-event, which are of keen interest in COVID-19 treatment trials. Moreover, even for binary outcomes, for which one possible adjustment method (Ge et al., 2011) was cited in the recent FDA guidance (FDA, 2020), there has not been any study showing how much precision gain is to be expected by using covariate adjusted, rather than unadjusted, methods in the context of COVID-19 treatment trials. In this work, we evaluate the performance of covariate adjusted estimators (hence, simply *adjusted estimators*) for binary, ordinal, and time-to-event outcomes.

To understand the idea, imagine repeating a trial many times with consideration of age as a key prognostic baseline variable. Due to chance, some trials will have more older participants assigned to treatment than control, and some will have more younger participants assigned to treatment than control. While the distribution of unadjusted estimates over these many trials will be centered at the true effect, it will be a mixture of under- and over- estimates. The variability of the estimates can be ascribed to the variability of the outcome within each age group *as well as* the variability in the balance of age between treatment and control. To understand this latter point, consider a setting where older adults are at higher risk for a negative outcome. In this situation, the trials where more older participants are assigned to the treatment, the treatment will tend to look worse than in trials where more younger participants were assigned to the treatment. Unadjusted estimates ignore the important information about imbalances with respect to age leading to greater variability. Adjusted estimates seek to minimize this source of variability in the estimation procedure thereby improving their efficiency relative to the unadjusted estimator (Jiang et al., 2019).

To make these ideas more precise, consider a randomized trial involving a single, binary-valued baseline variable (e.g., age over 85), two study arms (e.g., drug A vs. B), a binary outcome (e.g., death by 30 days after hospitalization), and the estimand being the risk difference (probability of death under assignment to treatment minus probability of death under assignment to control). The unadjusted estimator of the risk difference is the difference between arms in the sample proportion of patients who die. The adjusted estimator of the risk difference is constructed by first computing, separately for those > 85 and ≤ 85, the difference between arms in the sample proportions of patients who die; next, these stratum- specific treatment effect estimates are combined into an average treatment effect estimate by taking their weighted sum with weights set to the study-wide (pooling across arms) proportions of those > 85 and ≤ 85. Provided the sample size is large enough, the adjusted estimator will have greater precision compared to the unadjusted estimator if age > 85 is correlated with death (which is likely for COVID-19 positive patients). In general, the adjusted estimator can be interpreted as correcting for empirical confounding by baseline variables (Moore et al., 2011).

To clarify *how* the adjusted estimator gains information compared to the unadjusted estimator, consider the same setup as the previous paragraph except where the goal is even simpler: to estimate the probability of death under assignment to the treatment arm. The unadjusted estimator is the sample proportion of dead patients in the treatment arm. It can be equivalently represented as the weighted sum of stratum-specific sample proportions of dead patients with weights set to the proportions of those > 85 and ≤ 85 *in the treatment arm*. The adjusted estimator is the weighted sum of stratum-specific sample proportions of dead patients in the treatment arm with weights set to the *study-wide* (*pooling across arms*) *proportions of those >* 85 *and* ≤ 85. The only difference between the estimators is in the italicized text; the adjusted estimator uses a more precise estimate of the proportions in each stratum, which propagates into a more precise estimate of the probability of death under assignment to the treatment arm. A similar mechanism underlies the precision gains due to covariate adjustment when estimating the risk difference and the other estimands in this paper.

## 3. Estimands and Analysis Methods

Throughout, we assume that treatment is randomly assigned independently of baseline covariates. The described methods can be adapted to handle the case where treatment assignment depends on (a subset of) the measured baseline covariates, as is the case when stratified randomization is used. All estimands are intention-to-treat (ITT) in that they are contrasts between outcome distributions under assignment to treatment and under assignment to control. We let A denote study arm assignment, taking on the value 0 for control group and 1 for treatment group. We let Y denote the outcome of interest and X denote a vector of baseline covariates.

### 3.1 Binary Outcomes

We consider three estimands, though our simulation studies only involve the first. All probabilities below are marginal (as opposed to conditional on baseline variables). The outcome is coded as “good” (1) or “bad” (0).

Estimand 1: **Risk Difference**.

Difference between probability of bad outcome comparing treatment to control arms.

Estimand 2: **Relative Risk**.

Ratio of probability of bad outcome comparing treatment to control arms.

Estimand 3: **Odds Ratio**.

Ratio of odds of bad outcome, comparing treatment to control arms.

Estimators of each estimand 1–3 above can be constructed from estimators of the probability of a bad outcome for each study arm; e.g., the risk difference can be estimated by the difference between the arm-specific estimators. The unadjusted estimator of the probability of a bad outcome under assignment to each study arm *a* is just the sample proportion of bad outcomes among patients assigned to arm *A* = *a*. A covariate adjusted estimator of this quantity can be based on the standardization approach of Ge et al. (2011), as indicated in the FDA COVID-19 guidance (FDA, 2020). This estimator is identical to that of Moore and van der Laan (2009) and for the risk difference it is a special case of estimators from Scharfstein et al. (1999). First, a logistic regression model is fit for the probability of bad outcome given study arm and baseline variables. Next, for each participant (from both arms), a predicted probability of bad outcome is obtained under each possible arm assignment *a* ϵ {0,1} by plugging in the participant’s baseline variables and setting arm assignment A = 0 and A =1, respectively, in the logistic regression model fit. Lastly, the covariate adjusted estimator of the bad outcome probability in arm *a* ϵ {0,1} is the sample mean over all participants (pooling across arms) of the predicted outcome setting *A* = *a*.

### 3.2 Ordinal Outcomes

We consider three estimands when the outcome is ordinal, with levels 1,…,*K*. Without loss of generality, we assume that higher values of the ordinal outcome are preferable. We comment on these estimands briefly here, and refer the reader to Appendix A of the Supplementary Material for analytic expressions for these quantities.

Estimand 1: **Difference in means of a prespecified transformation of the ordinal outcome (DIM)**.

In most settings, this transformation will be monotone increasing so that larger values of the ordinal outcome will result in larger, and therefore preferable, transformed outcomes. Transformations could incorporate, e.g., utilities assigned to each level, as has been done in some stroke trials (Chaisinanunkul et al., 2015; Nogueira et al., 2018).

Estimand 2: **Mann-Whitney (MW) estimand**.

This estimand reports the probability that a random individual assigned to treatment will have a better outcome than a random individual assigned to control, with ties broken at random.

Estimand 3: **Log-odds ratio (LOR)**.

We consider a nonparametric extension of the log-odds ratio (LOR) (Díaz et al., 2016) defined as the average of the cumulative log odds ratios over levels 1 to K – 1 of the outcome. In the case that the distribution of the outcome given study arm is accurately described by a proportional odds model of the outcome against treatment (McCullagh, 1980), this estimand is equal to the coefficient associated with treatment.

As is shown in Appendix A of the Supplementary Material, all of the above estimands can be written as smooth summaries of the treatment-specific cumulative distribution functions (CDFs) of the ordinal outcome. To estimate these quantities, it suffices to estimate these two CDFs and then to evaluate the summaries. This results in so-called plug-in estimators.

The unadjusted estimator of the CDF in each arm is the empirical distribution in that arm. The resulting plug-in estimator for the DIM corresponds precisely to taking the difference of sample means of the transformed outcomes between the two arms. Also, the resulting plug-in estimator (denoted M) for the MW estimand is closely related to the usual Mann-Whitney U-statistic U = *n*_0_*n*_1_*M*, where *n*_0_ and *n*_1_ are the total sample sizes in the two study arms.

The covariate adjusted estimator of the CDF in each arm leverages prognostic information in baseline variables. It uses working models, i.e., models that are fit in the process of computing the estimator but which we do not assume to be correctly specified (and for which consistency and asymptotic normality still hold under arbitrary model misspecification). Specifically, the adjusted estimator of the CDF for each study arm is based on an arm-specific, proportional odds working model for the cumulative probability of the outcome given the baseline variables. This working model posits that, conditioned on arm *A* = *a* and a linear combination of the baseline variables, the odds of *Y* ≤ *j* is proportional to the odds of *Y* ≤ *j* + 1, where the proportionality constant depends on *j* but not on the baseline variables. Formally, the model for arm *A* = *a* is logit {*P*(*Y ≤ j* | *A* = *a, X*)} = *α_j_* + *β^T^X*, for each *j* = 1, …,*K* – 1 with parameters *α*_1_,…, *α*_K-1_ and *β*; the model for the other study arm is the same but with a separate set of parameters. Each model is fit using data from the corresponding study arm, yielding two working covariate-conditional CDFs (one per arm). For each arm, the estimated marginal CDF is then obtained by averaging the corresponding conditional CDF across the empirical distribution of baseline covariates pooled across the two study arms. This is analogous to standardizing arm-specific mean outcomes based on pooling baseline variables across arms as explained in the last paragraph of Section 2. The above methods are implemented in an accompanying R package, drord, available on GitHub at https://github.com/benkeser/drord.

The validity (i.e., consistency and asymptotic normality) of the adjusted CDF estimator given above in no way relies on correct specification of the aforementioned working model. The fact that our adjusted CDF estimator is robust to misspecification of the outcome working model is reminiscent of the fact that ANCOVA can be robustly used for covariate adjustment when the estimand is the difference in means between arms (Yang and Tsiatis, 2001; Tsiatis et al., 2008). Other model-robust, covariate adjusted estimators are available for estimation of the MW estimand, e.g., Vermeulen et al. (2015), and for the LOR estimand, e.g., Díaz et al. (2016).

### 3.3 Time-to-Event Outcomes

We consider three treatment effect estimands in the time-to-event setting, all of which are interpretable under violations of a proportional hazards assumption. We refer the reader to Appendix B of the Supplementary Material for analytic expressions for these quantities.

**Estimand 1: Difference in restricted mean survival times (RMSTs)**.

The RMST is the expected value of a survival time that is truncated at a specified time (Chen and Tsiatis, 2001; Royston and Parmar, 2011). Equivalently, the RMST can be expressed as the area under the survival curve up to the truncation time.

**Estimand 2: Survival probability difference (also called risk difference, RD)**

Difference between arm-specific probabilities of survival to a specified time.

**Estimand 3: Relative risk (RR)**.

Ratio of the arm-specific probabilities of survival to a specified time.

Analogous to the ordinal outcome case, estimators of these parameters can be constructed from estimators of the survival function at each time point. One approach to constructing adjusted estimators, used here, involves discretizing time and then: (i) estimating the time-specific hazard conditional on baseline variables, (ii) transforming to survival probabilities using the product-limit formula, and (iii) marginalizing using the estimated covariate distribution (pooled across arms). The adjusted approach as implemented here (and elsewhere-see references below) has two key benefits relative to unadjusted alternatives such as using the unadjusted Kaplan-Meier estimator (Kaplan and Meier, 1958). First, in large samples and under regularity conditions, the adjusted estimator is at least as precise as the unadjusted estimator. Second, the adjusted estimator’s consistency depends on an assumption of censoring being independent of the outcome given study arm and baseline covariates, rather than an assumption of censoring in each arm being independent of the outcome marginally; see Appendix B of the Supplementary Material for details. The former may be a more plausible assumption.

We implemented the covariate adjusted estimator of the RMST (specifically, the targeted minimum loss-based estimator of the RMST) from Díaz et al. (2019), which is implemented in the R package survtmlerct available on GitHub at https://github.com/idiazst/survtmlerct. Time was discretized as the day level. Similar covariate adjusted estimators for the RD and RR are also available (Moore and van der Laan, 2009; Benkeser et al., 2018, 2019). Both Díaz et al. (2019) and Benkeser et al. (2018) provide approaches that can be used to develop Wald-type confidence intervals and corresponding tests of the null hypothesis of no treatment effect.

Some examples of other covariate adjusted estimators for time-to-event outcomes include Stitelman et al. (2011); Brooks et al. (2013); Lu and Tsiatis (2011); (Zhang 2014) and Parast et al. (2014). Díaz et al. (2019) compare the properties of these estimators.

## 4. Simulation Methods

### 4.1 Sample sizes and number of simulated trials

In each setting, we simulated trials with 1:1 randomization to the two arms and total enrollment of *n* = 100, 200, 500, and 1000. In each case, 1000 trials were simulated.

### 4.2 Data-Generating Distributions

#### 4.2.1 Binary outcomes

The data generating distributions are the same as for ordinal outcomes (below), except that we dichotomized the outcome into “bad” (ICU admission or death) and “good” (neither ICU admission nor death).

#### 4.2.2 Ordinal outcomes

We generated data based on (CDC COVID-19 Response Team, 2020), which reported outcomes for hospitalized and non-hospitalized individuals with COVID- 19. We focus on hospitalized patients and place additional results pertaining to the nonhospitalized population in Appendix C of the Supplementary Material. There were three levels of the ordinal outcome, with level 1 representing death, level 2 representing survival with ICU admission, and level 3 representing survival without ICU admission. The following patient age categories define the single baseline variable (which is used for adjustment): 0-19, 20-44, 45-54, 55-64, 65-74, 75-84, and ≥85. Lower and upper estimates were given for probabilities of outcomes in each age group in (CDC COVID-19 Response Team, 2020); we used the mean of these within each age group to define our data generating distributions. For the hospitalized COVID-19 positive population, the resulting outcome probabilities for each age group are listed in Table 1.

**Table 1.** Hospitalized, COVID-19 positive population: Age and conditional outcome distributions based on data from (CDC COVID-19 Response Team, 2020) that we use for defining the control arm distribution in the ordinal outcome simulation studies. “ICU” represents ICU admission.

| Age | P(age) | P(death \| age) | P(ICU & survived \| age) | P(no ICU & survived \| age) |
| --- | --- | --- | --- | --- |
| 0–19 | 0.01 | 0.00 | 0.00 | 1.00 |
| 20–44 | 0.09 | 0.01 | 0.18 | 0.81 |
| 45–54 | 0.12 | 0.03 | 0.32 | 0.65 |
| 55–64 | 0.13 | 0.08 | 0.31 | 0.61 |
| 65–74 | 0.18 | 0.11 | 0.37 | 0.52 |
| 75–84 | 0.22 | 0.17 | 0.47 | 0.36 |
| $\geq 85$ | 0.25 | 0.37 | 0.35 | 0.28 |

We separately considered two types of treatment effects in our data generating distributions: no treatment effect and an effective treatment. For the former, we randomly sampled *n* age-outcome pairs according to the distribution in Table 1 and then independently assigned study arm with probability 1/2 for each arm.

For the latter case (effective treatment), we randomly generated control arm participants as in the previous paragraph. We randomly generated treatment arm participants using a modified version of Table 1 where the probabilities P(ICU admission and survived | age) in column 4 were proportionally reduced; then each corresponding probability in column 5, i.e., P(No ICU admission and survived | age), was increased by an equal amount. The probabilities of death given age in column 3 were not changed. This modified table corresponds to the treatment having no effect on the probability of death but decreasing the odds of ICU admission among those who survive by the same relative amount in each age category. This relative reduction (and the resulting treatment effect) was separately selected for each sample size *n* = 100, 200, 500, 1000.

In our simulations, we used the adjusted estimator described in Section 3.2, where age is coded using the categories in Table 1. Specifically, these age categories are included as the main terms in the linear parts of the proportional odds working models.

For simplicity, for binary and ordinal outcomes we simulated trials with no censoring. However, the methods we used can adjust for dropout (right censoring) as described in Appendix A of the Supplementary Material.

#### 4.2.3 Time-to-event outcomes

In order to mimic key predictive features of clinical outcomes in COVID patients, our simulation data generating mechanism is based on the over 500 patients hospitalized at Weill Cornell Medicine New York Presbyterian Hospital prior to March 28, 2020. The event of interest in this simulation is time from hospitalization to the first of intubation or death, and the predictive variables used are sex, age, whether the patient required supplemental oxygen at ED presentation, dyspnea, hypertension, and the presence of bilateral infiltrates on the chest x-ray. We focus on the RMST 14 days after hospitalization, and the RD of remaining intubation-free and alive 7 days after hospitalization.

Patient data was re-sampled with replacement to generate 1000 datasets of each of the sizes *n* = 100, 200, 500, and 1000. For each dataset, a hypothetical treatment variable was drawn from a Bernoulli distribution with probability 0.5 independently of all other variables. A positive treatment effect was simulated by adding an independent random draw from a *χ*^2^ distribution with 4 degrees of freedom to each participant’s outcome in the treatment arm. This corresponds to a difference in RMST of 1.06 at 14 days, and an RD of 8.7% at seven days. Five percent of the patients were selected at random to be censored, and their censoring time was drawn from a uniform distribution on {1,…, 14}.

We compare the performance of the unadjusted, Kaplan-Meier-based estimator to the covariate adjusted estimator defined in Sections 4 and 6 of (Díaz et al., 2019), respectively, and implemented in the R package survtmlerct. Wald-type confidence intervals and corresponding tests of the null hypothesis of no effect are reported.

### 4.3 Performance Criteria

We compare the type I error and power of tests of the null hypothesis *H*_0_ of no treatment effect based on unadjusted and adjusted estimators, both within and across estimands. For each estimand, we also compare the bias, variance, and mean squared error of the unadjusted and the adjusted estimators. We report the relative efficiency of the unadjusted relative to the adjusted estimator (ratio of variance of adjusted estimator to variance of unadjusted estimator). One minus this relative efficiency is approximately the proportion reduction in sample size needed for a covariate adjusted estimator to achieve the same power as the unadjusted estimator, asymptotically (van der Vaart, 1998, pp. 110-111).

## 5. Simulation Results

For binary and ordinal outcomes, we present results that use the nonparametric BCa bootstrap (Efron and Tibshirani, 1994) for confidence intervals and hypothesis tests. We used 1000 replicates for each BCa bootstrap confidence interval. While we recommend 10,000 replicates in practice, the associated computational time was too demanding for our simulation study. Nonetheless, we expect similar or slightly better performance with an increased number of bootstrap samples. Results that use closed-form, Wald-based inference methods are presented in Appendix C of the Supplementary Material.

For time-to-event outcomes, we used Wald-based confidence intervals since these made the computations faster compared to the BCa bootstrap method. For all outcome types and estimands, the relative efficiency of the unadjusted estimator to the adjusted estimator is approximated by the ratio of the mean squared error of the latter to the mean squared error of the former.

### 5.1 Binary Outcomes

Table 2 compares the performance of the unadjusted and adjusted estimators when the outcome is ICU admission or death, and the estimand is the risk difference. Type I error of the covariate adjusted method was comparable to that of the unadjusted method. The covariate adjusted method achieved higher power across all settings. Absolute gains in power varied from 5% to 12%.

The relative efficiency of unadjusted method relative to the adjusted method varied from 0.90 to 0.84. This is roughly equivalent to needing 10-16% smaller sample size when using the adjusted estimator compared to the unadjusted estimator, to achieve the same power.

**Table 2.** Results for the binary outcome and risk difference (RD) estimand in the hospitalized population. BCa bootstrap is used for confidence intervals and hypothesis testing. “Effect” denotes the true estimand value; “MSE” denotes mean squared error; “Rel. Eff.” denotes relative efficiency which we approximate as the ratio of the MSE of the estimator under consideration to the MSE of the unadjusted estimator. In each block of four rows, the first two rows involve no treatment effect and the last two rows involve a benefit from treatment.

| $n$ | Estimator Type | Effect | P(reject $H_0$) | MSE | Bias | Variance | Rel. Eff. |
| --- | --- | --- | --- | --- | --- | --- | --- |
| 100 | Unadjusted | 0 | 0.043 | 0.010 | 0.003 | 0.010 | 1.000 |
| 100 | Adjusted | 0 | 0.049 | 0.009 | 0.004 | 0.009 | 0.844 |
| 100 | Unadjusted | -0.269 | 0.719 | 0.009 | 0.003 | 0.009 | 1.000 |
| 100 | Adjusted | -0.269 | 0.841 | 0.008 | 0.004 | 0.008 | 0.859 |
| 200 | Unadjusted | 0 | 0.031 | 0.005 | 0.003 | 0.005 | 1.000 |
| 200 | Adjusted | 0 | 0.043 | 0.004 | 0.004 | 0.004 | 0.885 |
| 200 | Unadjusted | -0.199 | 0.768 | 0.005 | 0.003 | 0.005 | 1.000 |
| 200 | Adjusted | -0.199 | 0.835 | 0.004 | 0.004 | 0.004 | 0.880 |
| 500 | Unadjusted | 0 | 0.047 | 0.002 | 0.001 | 0.002 | 1.000 |
| 500 | Adjusted | 0 | 0.047 | 0.002 | 0.000 | 0.002 | 0.878 |
| 500 | Unadjusted | -0.124 | 0.770 | 0.002 | 0.000 | 0.002 | 1.000 |
| 500 | Adjusted | -0.124 | 0.836 | 0.002 | 0.000 | 0.002 | 0.899 |
| 1000 | Unadjusted | 0 | 0.041 | 0.001 | 0.000 | 0.001 | 1.000 |
| 1000 | Adjusted | 0 | 0.045 | 0.001 | 0.000 | 0.001 | 0.860 |
| 1000 | Unadjusted | -0.090 | 0.796 | 0.001 | 0.000 | 0.001 | 1.000 |
| 1000 | Adjusted | -0.090 | 0.852 | 0.001 | 0.000 | 0.001 | 0.890 |

### 5.2 Ordinal outcomes

Tables 3, 4, and 5 display results for the difference in means (DIM), Mann-Whitney (MW), and log odds ratio (LOR) estimands, respectively. Type I error control of the covariate adjusted methods was comparable to that of the unadjusted methods. The covariate adjusted methods achieved higher power across all settings. Absolute gains in power varied from 3% to 8% for the DIM, 1% to 6% for the MW estimand, and 3% to 6% for the LOR.

The relative efficiency of the unadjusted methods relative to adjusted methods varied from 0.89 to 0.82 for the DIM, 0.91 to 0.82 for the MW estimand, and 0.88 to 0.82 for the LOR. This is roughly equivalent to needing 11-18% (DIM), 9-18% (MW), and 12-18% (LOR) smaller sample sizes, respectively, when using the adjusted estimator compared to the unadjusted estimator, to achieve the same power.

**Table 3.** Results for the ordinal outcome and difference in means (DIM) estimand in the hospitalized population. BCa bootstrap is used for confidence intervals and hypothesis testing. “Effect” denotes the true estimand value; “MSE” denotes mean squared error; “Rel. Eff.” denotes relative efficiency which we approximate as the ratio of the MSE of the estimator under consideration to the MSE of the unadjusted estimator. In each block of four rows, the first two rows involve no treatment effect and the last two rows involve a benefit from treatment.

| $n$ | Estimator Type | Effect | P(reject $H_0$) | MSE | Bias | Variance | Rel. Eff. |
| --- | --- | --- | --- | --- | --- | --- | --- |
| 100 | Unadjusted | 0 | 0.064 | 0.023 | -0.005 | 0.023 | 1.000 |
| 100 | Adjusted | 0 | 0.058 | 0.019 | -0.007 | 0.019 | 0.822 |
| 100 | Unadjusted | 0.303 | 0.472 | 0.022 | -0.007 | 0.022 | 1.000 |
| 100 | Adjusted | 0.303 | 0.553 | 0.019 | -0.004 | 0.019 | 0.845 |
| 200 | Unadjusted | 0 | 0.049 | 0.010 | -0.002 | 0.010 | 1.000 |
| 200 | Adjusted | 0 | 0.045 | 0.009 | -0.003 | 0.009 | 0.862 |
| 200 | Unadjusted | 0.303 | 0.775 | 0.012 | -0.003 | 0.012 | 1.000 |
| 200 | Adjusted | 0.303 | 0.842 | 0.010 | 0.000 | 0.010 | 0.872 |
| 500 | Unadjusted | 0 | 0.061 | 0.005 | -0.001 | 0.005 | 1.000 |
| 500 | Adjusted | 0 | 0.057 | 0.004 | 0.000 | 0.004 | 0.837 |
| 500 | Unadjusted | 0.195 | 0.810 | 0.005 | 0.000 | 0.005 | 1.000 |
| 500 | Adjusted | 0.195 | 0.855 | 0.004 | 0.001 | 0.004 | 0.891 |
| 1000 | Unadjusted | 0 | 0.052 | 0.002 | 0.000 | 0.002 | 1.000 |
| 1000 | Adjusted | 0 | 0.042 | 0.002 | 0.000 | 0.002 | 0.849 |
| 1000 | Unadjusted | 0.136 | 0.835 | 0.002 | 0.000 | 0.002 | 1.000 |
| 1000 | Adjusted | 0.136 | 0.867 | 0.002 | 0.000 | 0.002 | 0.889 |

**Table 4.** Results for ordinal outcome and Mann Whitney (MW) estimand in the hospitalized population. BCa bootstrap is used for confidence intervals and hypothesis testing. “Effect” denotes the true estimand value; “MSE” denotes mean squared error; “Rel. Eff.” denotes relative efficiency which we approximate as the ratio of the MSE of the estimator under consideration to the MSE of the unadjusted estimator. In each block of four rows, the first two rows involve no treatment effect and the last two rows involve a benefit from treatment.

| $n$ | Estimator Type | Effect | P(reject $H_0$) | MSE | Bias | Variance | Rel. Eff. |
| --- | --- | --- | --- | --- | --- | --- | --- |
| 100 | Unadjusted | 0.500 | 0.054 | 0.003 | -0.002 | 0.003 | 1.000 |
| 100 | Adjusted | 0.500 | 0.055 | 0.002 | -0.003 | 0.002 | 0.822 |
| 100 | Unadjusted | 0.627 | 0.688 | 0.002 | -0.002 | 0.002 | 1.000 |
| 100 | Adjusted | 0.627 | 0.753 | 0.002 | -0.002 | 0.002 | 0.852 |
| 200 | Unadjusted | 0.500 | 0.047 | 0.001 | -0.001 | 0.001 | 1.000 |
| 200 | Adjusted | 0.500 | 0.036 | 0.001 | -0.001 | 0.001 | 0.864 |
| 200 | Unadjusted | 0.627 | 0.940 | 0.001 | -0.001 | 0.001 | 1.000 |
| 200 | Adjusted | 0.627 | 0.968 | 0.001 | 0.000 | 0.001 | 0.878 |
| 500 | Unadjusted | 0.500 | 0.055 | 0.001 | 0.000 | 0.001 | 1.000 |
| 500 | Adjusted | 0.500 | 0.052 | 0.000 | 0.000 | 0.000 | 0.843 |
| 500 | Unadjusted | 0.582 | 0.946 | 0.001 | 0.000 | 0.001 | 1.000 |
| 500 | Adjusted | 0.582 | 0.959 | 0.000 | 0.000 | 0.000 | 0.905 |
| 1000 | Unadjusted | 0.500 | 0.046 | 0.000 | 0.000 | 0.000 | 1.000 |
| 1000 | Adjusted | 0.500 | 0.046 | 0.000 | 0.000 | 0.000 | 0.844 |
| 1000 | Unadjusted | 0.557 | 0.936 | 0.000 | 0.000 | 0.000 | 1.000 |
| 1000 | Adjusted | 0.557 | 0.948 | 0.000 | 0.000 | 0.000 | 0.890 |

**Table 5.**
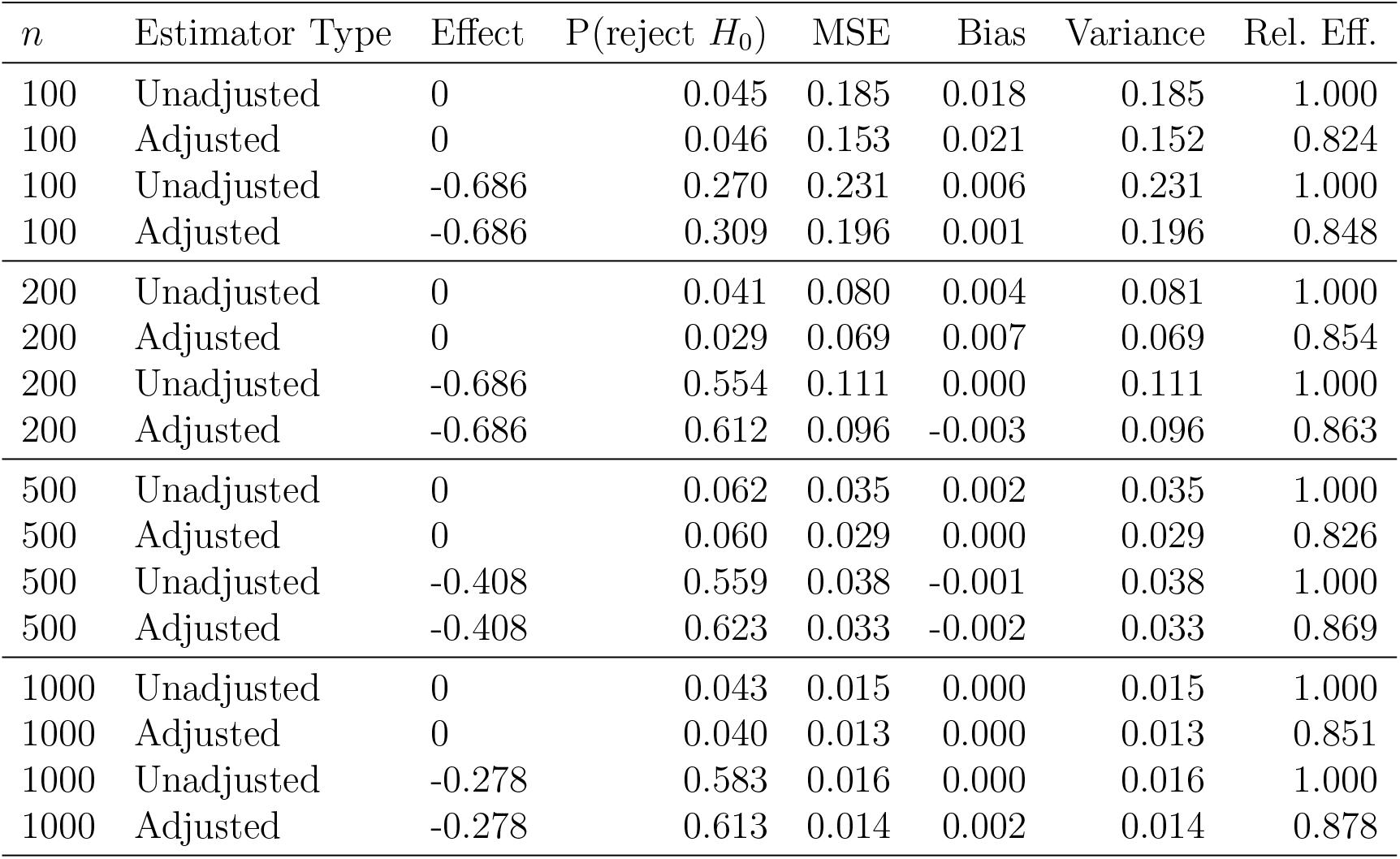
Results for the ordinal outcome and log-odds ratio (LOR) estimand in the hospitalized population. BCa bootstrap is used for confidence intervals and hypothesis testing. “Effect” denotes the true estimand value; “MSE” denotes mean squared error; “Rel. Eff.” denotes relative efficiency which we approximate as the ratio of the MSE of the estimator under consideration to the MSE of the unadjusted estimator. In each block of four rows, the first two rows involve no treatment effect and the last two rows involve a benefit from treatment.

| $n$ | Estimator Type | Effect | P(reject $H_0$) | MSE | Bias | Variance | Rel. Eff. |
| --- | --- | --- | --- | --- | --- | --- | --- |
| 100 | Unadjusted | 0 | 0.045 | 0.185 | 0.018 | 0.185 | 1.000 |
| 100 | Adjusted | 0 | 0.046 | 0.153 | 0.021 | 0.152 | 0.824 |
| 100 | Unadjusted | -0.686 | 0.270 | 0.231 | 0.006 | 0.231 | 1.000 |
| 100 | Adjusted | -0.686 | 0.309 | 0.196 | 0.001 | 0.196 | 0.848 |
| 200 | Unadjusted | 0 | 0.041 | 0.080 | 0.004 | 0.081 | 1.000 |
| 200 | Adjusted | 0 | 0.029 | 0.069 | 0.007 | 0.069 | 0.854 |
| 200 | Unadjusted | -0.686 | 0.554 | 0.111 | 0.000 | 0.111 | 1.000 |
| 200 | Adjusted | -0.686 | 0.612 | 0.096 | -0.003 | 0.096 | 0.863 |
| 500 | Unadjusted | 0 | 0.062 | 0.035 | 0.002 | 0.035 | 1.000 |
| 500 | Adjusted | 0 | 0.060 | 0.029 | 0.000 | 0.029 | 0.826 |
| 500 | Unadjusted | -0.408 | 0.559 | 0.038 | -0.001 | 0.038 | 1.000 |
| 500 | Adjusted | -0.408 | 0.623 | 0.033 | -0.002 | 0.033 | 0.869 |
| 1000 | Unadjusted | 0 | 0.043 | 0.015 | 0.000 | 0.015 | 1.000 |
| 1000 | Adjusted | 0 | 0.040 | 0.013 | 0.000 | 0.013 | 0.851 |
| 1000 | Unadjusted | -0.278 | 0.583 | 0.016 | 0.000 | 0.016 | 1.000 |
| 1000 | Adjusted | -0.278 | 0.613 | 0.014 | 0.002 | 0.014 | 0.878 |

### 5.3 Time-to-event outcomes

Table 6 displays the results for RMST estimators, where the baseline variables adjusted for include age and sex along with the four other variables described in Section 4.2.3. Type I error control of the covariate adjusted method was comparable to that of the unadjusted method. The covariate adjusted methods achieved higher power across all settings, with absolute gains varying from 3% to 8%. The value added by covariate adjustment in terms of relative efficiency increased monotonically as a function of sample size, from approximately 0.95 (roughly equivalent to needing 5% smaller sample size) when *n* = 100 to approximately 0.79 (roughly equivalent to needing 21% smaller sample size) when *n* = 1000 for the case of no treatment effect; the results were similar for the positive treatment effect case, except with slightly smaller precision gains.

To evaluate the importance of adjusting for multiple baseline variables, we also evaluated an adjusted RMST estimator that only adjusts for age and sex in Appendix C of the Supplementary Material. The gains of the covariate adjusted methods relative to the unadjusted methods were more modest, with absolute gains in power of approximately 0%- 1% and relative efficiency ranging from 0.96 to 1.00. These results suggest that there can be a meaningful benefit from adjusting for prognostic covariates beyond just age and sex.

We also considered the risk difference (RD) estimand in Appendix C of the Supplementary Material. The results (when adjusting for age and sex along with the four other variables described in Section 4.2.3) are qualitatively similar, except with slightly smaller precision gains, to those for the RMST in Table 6.

**Table 6.** Results for difference in restricted mean survival times (RMST) at 14 days estimand in hospitalized population, when the adjusted estimator uses all six baseline variables from Section 4.2.3. Confidence intervals and hypothesis tests are Wald-based. “Effect” denotes the true estimand value; “MSE” denotes mean squared error; “Rel. Eff.” denotes relative efficiency which we approximate as the ratio of the MSE of the estimator under consideration to the MSE of the unadjusted estimator. In each block of four rows, the first two rows involve no treatment effect and the last two rows involve a benefit from treatment.

| Sample Size | Estimator Type | Effect | P(reject $H_0$) | MSE | Bias | Variance | Rel. Eff. |
| --- | --- | --- | --- | --- | --- | --- | --- |
| 100 | Unadjusted | 0 | 0.011 | 0.781 | 0.018 | 0.780 | 1.000 |
| 100 | Adjusted | 0 | 0.038 | 0.740 | 0.006 | 0.740 | 0.948 |
| 100 | Unadjusted | 1.06 | 0.085 | 0.570 | -0.265 | 0.500 | 1.000 |
| 100 | Adjusted | 1.06 | 0.160 | 0.546 | -0.251 | 0.483 | 0.957 |
| 200 | Unadjusted | 0 | 0.048 | 0.481 | -0.013 | 0.481 | 1.000 |
| 200 | Adjusted | 0 | 0.055 | 0.418 | -0.013 | 0.418 | 0.869 |
| 200 | Unadjusted | 1.06 | 0.326 | 0.328 | -0.145 | 0.307 | 1.000 |
| 200 | Adjusted | 1.06 | 0.397 | 0.293 | -0.123 | 0.278 | 0.893 |
| 500 | Unadjusted | 0 | 0.050 | 0.201 | -0.003 | 0.201 | 1.000 |
| 500 | Adjusted | 0 | 0.052 | 0.164 | -0.005 | 0.164 | 0.814 |
| 500 | Unadjusted | 1.06 | 0.729 | 0.151 | -0.070 | 0.146 | 1.000 |
| 500 | Adjusted | 1.06 | 0.810 | 0.129 | -0.065 | 0.125 | 0.855 |
| 1000 | Unadjusted | 0 | 0.048 | 0.100 | 0.001 | 0.100 | 1.000 |
| 1000 | Adjusted | 0 | 0.045 | 0.079 | 0.001 | 0.079 | 0.793 |
| 1000 | Unadjusted | 1.06 | 0.959 | 0.079 | -0.060 | 0.076 | 1.000 |
| 1000 | Adjusted | 1.06 | 0.986 | 0.065 | -0.058 | 0.061 | 0.819 |

## 6. Recommendations for Target of Inference and Primary Efficacy Analysis

1. **Estimand when the outcome is ordinal**. If a utility function can be agreed upon to transform the outcome to a score with a clinically meaningful scale, then we recommend using the difference between the transformed means in the treatment and control arms. Otherwise, we recommend using the unweighted difference between means or the Mann-Whitney estimand. We recommend against estimating log odds ratios, since these are difficult to interpret and the corresponding estimators (even unadjusted ones) can be unstable at small sample sizes; this instability occurrred in additional simulation studies that we describe in Appendix C of the Supplementary Material.
2. **Covariate adjustment**. Adjust for prognostic baseline variables to improve precision and power. We expect improvements to be substantial since there are already several known prognostic baseline variables, e.g., age and co-morbidities. The baseline variables should be specified before the trial is started (or should be selected using only blinded data from the trial using a prespecified algorithm). The number of variables adjusted for should be no more than (roughly) the total sample size divided by 20, as a rule of thumb to avoid model overfit.
3. **Confidence intervals and hypothesis testing**. We recommend that the nonparametric bootstrap (BCa method) be used with 10000 replicates for constructing a confidence interval. The entire estimation procedure, including any model fitting, should be repeated in each replicate data set. Hypothesis tests can be conducted either by inverting the confidence interval or by permutation methods – the latter can be especially useful in smaller sample size trials in order to achieve the desired Type I error rate. Vermeulen et al. (2015) present such a permutation-based test for the MW estimand based on a different covariate adjusted estimator than presented here.
4. **Early stopping for efficacy or futility**. Rules for early stopping such as O’Brien-Fleming boundaries can be directly applied, where z-statistics are constructed using the covariate adjusted estimator described above and the covariance between statistics at different analysis times is estimated using nonparametric bootstrap as described above. The timing of analyses (including the final analysis) can be based on the information accrued defined as the reciprocal of the adjusted estimator’s variance. In this way, precision gains from covariate adjustment translate into faster information accrual and shorter trial duration, even in trials with no treatment effect.
5. **Plotting the CDF and the probability mass function (PMF) when the outcome is ordinal**. Regardless of which treatment effect definition is used in the primary efficacy analysis, we recommend that the covariate adjusted estimate of the PMF and/or CDF of the primary outcome be plotted for each study arm when the outcome is ordinal. Pointwise and simultaneous confidence intervals should be displayed (where the latter account for multiple comparisons). This is analogous to plotting Kaplan-Meier curves for time-to-event outcomes, which can help in interpreting the trial results. For example, Figure 1 shows covariate adjusted estimates of the CDF and PMF for a data set from our simulation study. From the plots, it is evident that the effect of the treatment on the ordinal outcome is primarily through preventing ICU admission, with no impact on probability of death.
6. **Missing covariates**. We recommend handling missing covariates by imputing them based only on data from those covariates that were observed. Importantly, to ensure that treatment assignment is randomized conditionally on the imputed covariates, no treatment or outcome information should be used in this imputation.
7. **Missing ordinal outcomes**. We recommend handling missing ordinal outcomes using doubly robust methods whose validity relies on the outcomes being missing at random conditional on the covariates and study arm assignment. One such doubly robust approach can be evaluated by applying the methods described in Appendix A.2 of the Supplementary Material.
8. **Loss to follow up with time-to-event outcomes**. We recommend accounting for loss-to-follow-up using doubly robust methods such as those described in Benkeser et al. (2018); Díaz et al. (2019). These methods rely on a potentially more plausible condition on the censoring distribution than do unadjusted methods. The covariate adjusted estimator for the restricted mean survival time in the time-to-event setting is double robust under censoring being independent of the outcome given baseline variables and arm assignment.

## 7. Discussion

Alleviating the impact of the COVID-19 pandemic will likely require the development of effective interventions for treatments, as well as a preventive vaccine. In both cases, there is a pressing need to bring products to market as quickly as possible without sacrificing the validity of the analysis of trials used to evaluate therapeutics. Covariate adjustment represents a straightforward means of significantly improving the conduct of these trials, by more efficiently using data that are already routinely collected in the course of randomized trials. This in turn will allow effective treatments to be discovered with greater power and also end trials of ineffective treatments sooner, so that resources can be appropriately reallocated.

Of the phase 2 and 3 trials of treatments for COVID-19 that are registered on clinicaltrials.gov, there are currently 190 with target sample size above 200 (the sample size above which substantial precision gains were consistently observed in our simulation studies). Covariate adjustment may add value in these and future trials. Whether covariate adjustment is useful depends on how prognostic the baseline variables are for the primary outcome. Based on the moderate to strong correlations that drove the precision gains in our simulation studies, we expect that many COVID-19 trials will gain precision by adjusting for baseline variables.

Adjusting for baseline variables beyond just age and sex led to substantial improvements in precision in our simulations involving time-to-event outcomes. Specifically, for the simulated time-to-event intubation or death outcome among hospitalized patients at Weill Cornell Medicine, we saw meaningful gains from additionally adjusting for hypertension, dyspnea, whether the patient required supplemental oxygen at ED presentation, and the presence of bilateral infiltrates on the chest x-ray. For the other outcome types, i.e., binary and ordinal, our data generating distributions only had one baseline variable, age; this is all that was available in the CDC data, so we were not able to investigate the value added by adjusting for more variables.

Vermeulen et al. (2015) derived an adjusted estimator that is directly targeted at maximizing precision for the MW estimand. In contrast, our adjusted estimators for ordinal outcomes target the entire treatment-specific CDFs. It is an open research question to compare our methods to those of Vermeulen et al. (2015).

**Figure 1.**
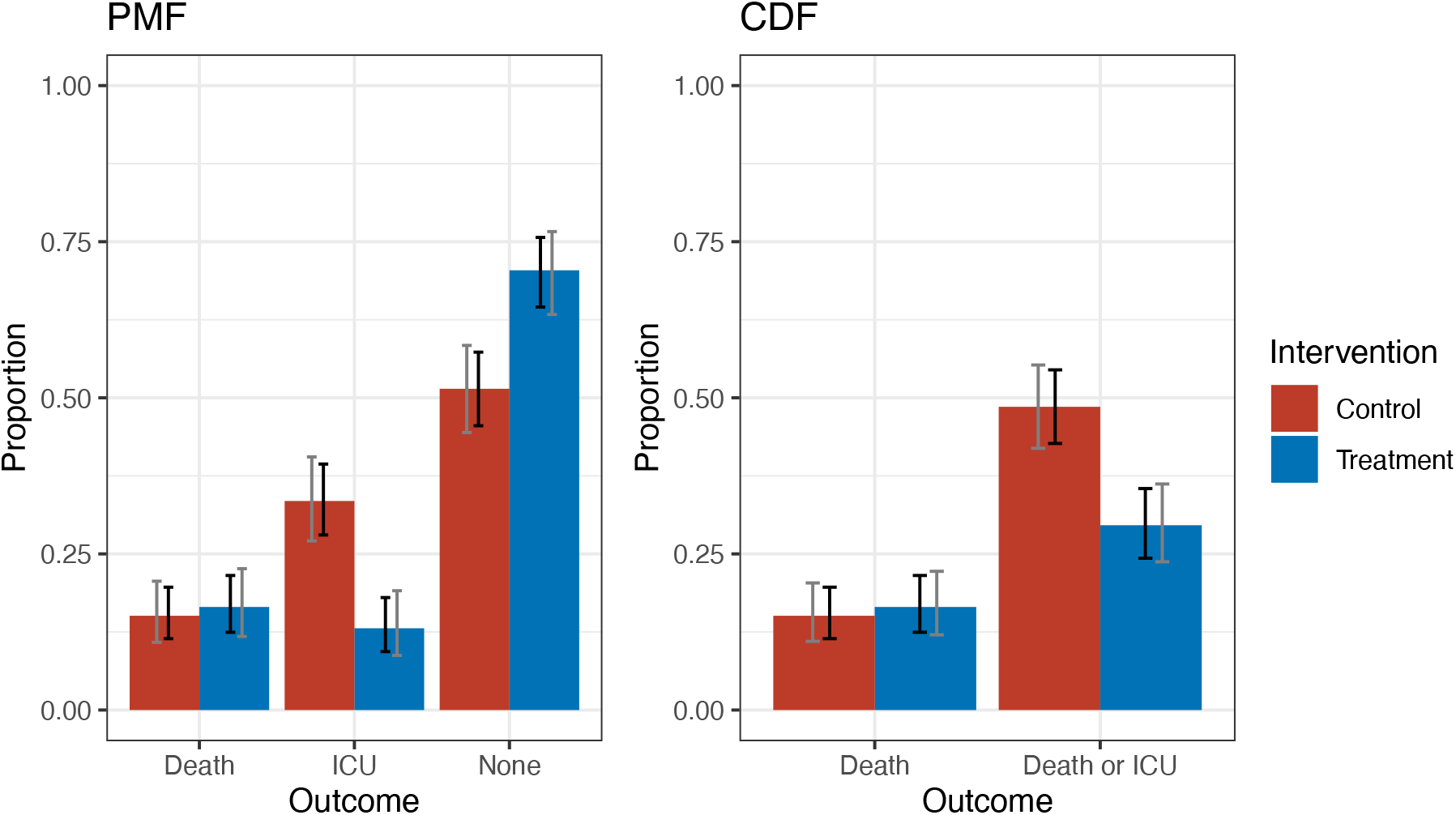
Example figures illustrating covariate adjusted estimates of the CDF and PMF by study arm with pointwise (black) and simultaneous (gray) confidence intervals. “ICU” represents survival and ICU admission; “None” represents survival and no ICU admission.

## Data Availability

The patient level data used to inform our simulations is not available to be shared.

## Supplementary Materials

Appendix A defines the estimands and estimators for ordinal outcomes. Appendix B describes the estimands and assumptions on censoring that we make for time-to-event outcomes. Appendix C presents additional simulation studies, including data generating distributions for non-hospitalized COVID-19 patients. Appendix D describes the availability of code that reproduces our simulation experiments and that implements our estimators and confidence intervals.

## Acknowledgements

AL was supported by the National Institutes of Health under award number DP2-LM013340. MR was supported by the Johns Hopkins Center of Excellence in Regulatory Science and Innovation funded by the U.S. Food and Drug Administration (FDA) U01FD005942. The content is solely the responsibility of the authors and does not necessarily represent the official views of the aforementioned organizations.

## Web Supplement

Summary: The Web Supplement is organized as follows. Appendix A introduces the estimands and estimators for ordinal outcomes. Appendix B introduces the estimands and assumptions on censoring that we make for time-to-event outcomes. Appendix C presents additional simulation studies, including for non-hospitalized COVID-19 patients. Appendix C.1 presents the data-generating distributions for non-hospitalized COVID-19 patients. Appendix C.2 presents the results of simulation studies for the case that the outcome is binary. Appendix C.3 presents additional simulation results for ordinal outcomes, namely the results for Wald-style inference and for the non-hospitalized population. Appendix C.4 presents additional simulation results for time-to-event outcomes, namely when a restricted set of covariates (age and sex) were used for adjustment and for the difference of survival probabilities in the hospitalized population. Appendix D describes the availability of code that reproduces our simulation experiments and that implements our estimator and confidence intervals.

## A. Estimands and estimators when the outcome is ordinal

### A.1 Estimands

Let (*A*, *Y*) and (*Ã*, *Ỹ*) denote independent treatment-outcome pairs, and let *u*(·) be a prespecified, real-valued transformation of an outcome. The three estimands are defined as follows:

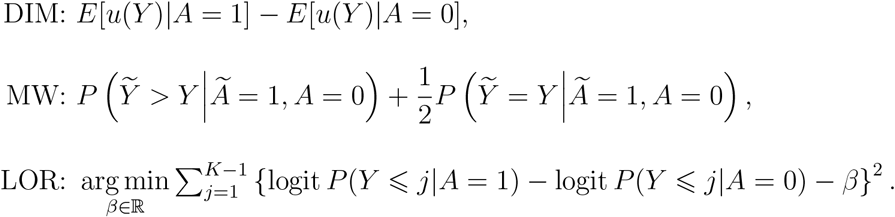

All three estimands are smooth summaries of the cumulative distribution functions *F_a_* (·):= *P*(*Y* ≤ · | *A* = *a*) for *a* 2 {0,1}. To see that this is the case, let *f_a_*(*j*):= *F_a_*(*j*) *– F_a_*(*j* – 1), *a ϵ* {0,1}, denote the corresponding probability mass functions and note that the estimands can be equivalently expressed as follows:

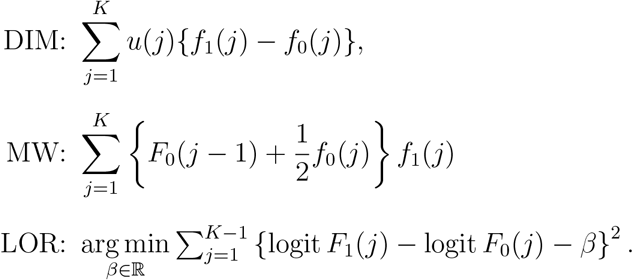

### A.2 Covariate adjusted estimator

Consider a setting in which we observe *n* independent copies of (*X, A*, *Y*), where *X* represents a *d*-dimensional vector of baseline covariates, *A* represents treatment, and *Y* represents outcome. We assume that *A* ╨ *X*. We use the subscript *i* to denote data specific to individual *i*. We now derive an estimator for the CDF that is closely related to an estimator presented in Scharfstein et al. (1999 ) and to targeted minimum loss-based estimators (van der Laan and Rubin 2006 van der Laan and Rose 2011).

For 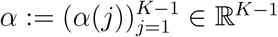 and *β* 2 ℝ*^d^*, define the following ℝ*^K^*^-1^ × ℝ*^d^* function:

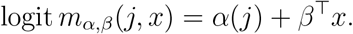

We will consider the treatment-stratified proportional odds working model for *P*{*Y* ≤ *j| A* = *a*, *X* = *x*} in which there exist (*α*_0_,*β*_0_) and (*α*_1_,*β*_1_) such that 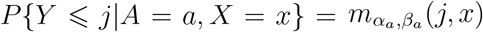 for all *j*,*x*, *a*. Importantly, we do not rely on this model being correct.

In addition to the above working model, we consider a treatment-assignment propensity score working model. It is used to define inverse-probability weights that are used when fitting the aforementioned proportional odds working models. Let 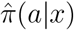 be an estimate of *P*(*A* = *a*|*X* = *x*), e.g., using a logistic regression model. In the clinical trial setting that we considered in our simulation studies, we used a logistic regression model with just an intercept, i.e., we ignored baseline variables. This is equivalent to using no weights (i.e., all weights equal to a constant) when fitting the proportional odds models. At the end of this subsection, we describe alternative approaches for estimating *P*(*A* = *a*|*X* = *x*) and the implications of doing so.

Suppose that, for *a ϵ* {0,1}, 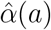 and 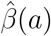 are chosen to minimize the following weighted empirical risk in (*α*, *β*):

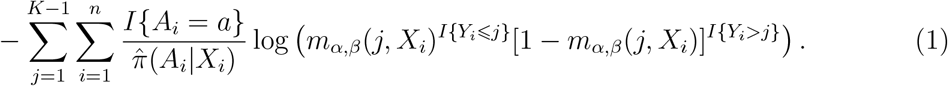

Each of these a-specific optimizations can be solved by running a weighted logistic regression on a repeated measures dataset of size *n* × (*K* – 1). Alternatively, they can be fit using software for a proportional odds model that allows for weights. For both levels of the treatment *a*, it can be shown that 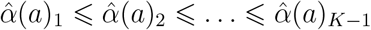, and so, for any covariate value *x*, 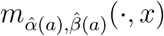 is a monotone nondecreasing function. Moreover, if our treatment-stratified proportional odds working model is correct, then 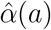 and 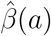 are consistent and asymptotically normal estimators of the true underlying parameters.

Our covariate adjusted estimate of the CDF *ψ_a_*(*j*):= *P*(*Y* ≤ *j* | *A* = *a*) is given by

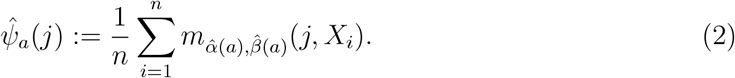

Because 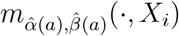 is monotone nondecreasing for all *i* = 1,…,*n*, *ψ_a_*(·) is also monotone nondecreasing. The above estimator also satisfies the known constraint that *ψ_a_*(*j*) ϵ [0,1].

It can also be shown that 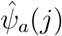 is (i) doubly robust and (ii) efficient if both the treatment mechanism (*P*(*A* = *a* |*X*)) working model and the stratified proportional odds working model are correctly specified. To show this, we now establish that 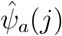 is in fact an augmented inverse probability weighted estimator. First, note that minimizing (1) to find 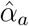 and 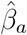 implies that the following first-order condition is satisfied for all *j* ϵ {1,…,*K* – 1}:

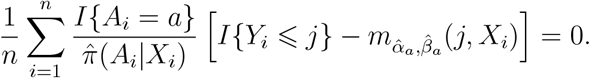

Next, note that adding this to the right-hand side of (2) shows that

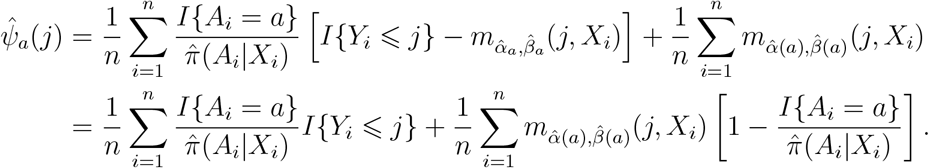

The above shows that 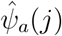 is an augmented inverse probability weighted estimator (see Section 7 of Robins et al. 1994) for 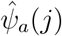, with the estimate of the outcome regression 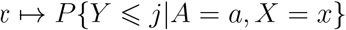 given by 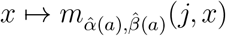.

We next discuss our estimation of the treatment probability *P*(*A* = *a* |*X* = *x*). Though this quantity can always be estimated by the empirical treatment probability in our randomized trial setting, there are generally advantages to estimating this quantity within a richer model. For example, a logistic regression of treatment on covariates (main effects only) could be used – in a randomized trial setting, this model is correctly specified provided that it includes an intercept term. The advantage of estimating known treatment probabilities via correctly specified parametric models has been discussed elsewhere - see, for example, Williamson et al. (2014) or, for a general treatment, Section 2.3.7 in van der Laan et al. (2003)

We recommend handling missing ordinal outcomes using doubly robust methods whose validity relies on the outcomes being missing at random conditional on the covariates and treatment assignment. To implement this approach, one can apply the methods described above, but with study arm recoded as 0 to indicate that a patient was both randomized to study arm 0 (control) and had their outcome measured, 1 to indicate that a patient was both randomized to study arm 1 (treatment) and had their outcome measured, and -1 to indicate that the outcome is missing. When study arm is recoded in this way and the outcome is not missing completely at random but is missing at random conditional on covariates, it is important that the model used for 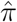 described above conditions on the baseline covariates, since this recoded treatment is not fully randomized.

## B. Estimands and censoring assumptions for time-to-event outcomes

Let *T* be a time-to-event outcome, *C* be a right-censoring time, *A* be a treatment indicator, and *X* be a collection of baseline covariates. Let τ be an investigator-specified truncation time that will be used to define the RMST, and let *t*\* be an investigator-specified time at which a comparison between the arm-specific survival probabilities is of interest.

The three estimands are defined as

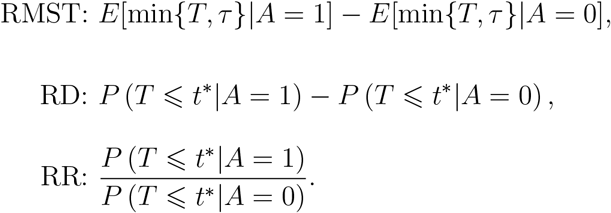

Unadjusted methods assume that

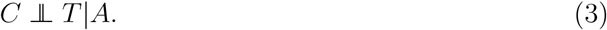

The adjusted methods discussed in the main text assume that

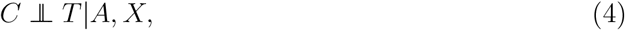

which may be more plausible than (3).

## C. Additional simulation studies

### C.1 Data generating distributions for non-hospitalized, COVID-19 patients

We also conducted simulations to mimic a population of non-hospitalized individuals who test positive for COVID-19 and where the primary outcome is ordinal (1=death, 2=hospitalized and survived, 3=not hospitalized and survived) and the baseline covariate is age category. We set the control arm probabilities of being in each age group and of hospitalization and death as in Table 1, which was extracted from CDC COVID-19 Response Team (2020) analogous to how this was done in Section 4.2.2 for the hospitalized population; the treatment arm distribution was constructed similarly as in Section 4.2.2.

Analogous to the hospitalized population data generating distributions, we assumed that a treatment would have no effect on the probability of death but would decrease the odds of hospital admission (hospitalization) by the same relative amount in each age category. For ordinal outcome scenarios with smaller sample sizes, there were sometimes data sets that had no participants in the lowest or highest outcome category in at least one study arm. For these data sets, the log-odds ratio estimators are undefined. As such, we omitted these sample sizes from our evaluations.

**Table 1.** Non-hospitalized,COVID-19 positive population: Age and conditional outcome distributions based on data from (CDC COVID-19 Response Team, 2020) that we use for defining the control arm distribution in the ordinal outcome simulation studies for the **non-hospitalized** population. “Hosp.” abbreviates “hospitalized”; “surv.” abbreviates “survived”.

| Age | P(age) | P(death \| age) | P(hosp. & surv. \| age) | P(not hosp. & surv. \| age) |
| --- | --- | --- | --- | --- |
| 0–19 | 0.05 | 0.00 | 0.02 | 0.98 |
| 20–44 | 0.29 | 0.00 | 0.18 | 0.82 |
| 45–54 | 0.18 | 0.01 | 0.25 | 0.74 |
| 55–64 | 0.18 | 0.02 | 0.25 | 0.73 |
| 65–74 | 0.17 | 0.04 | 0.36 | 0.60 |
| 75–84 | 0.09 | 0.07 | 0.45 | 0.48 |
| $\geq 85$ | 0.06 | 0.19 | 0.51 | 0.30 |

### C.2 Additional simulation studies for binary outcomes

We repeated the simulation studies in hospitalized and non-hospitalized patients for ordinal outcomes, but collapsing the death and ICU admission outcomes (hospitalized setting) and the death and hospitalized outcomes (non-hospitalized setting) to make a binary composite outcome. The binary outcome in the non-hospitalized population is defined as death or hospitalization (*Y* = 0) or survived and no hospitalization (*Y* = 1). The binary outcome for the hospitalized population is as defined in Section 4.2.1 of the main paper. We compared covariate-adjusted vs. unadjusted estimates of the risk difference of the binary outcome in terms of mean squared error, bias, and variance. We also compared the probability of rejecting the null hypothesis of 0 risk difference using a test based on our covariate-adjusted estimator versus a traditional Chi-squared test. Results are shown in Tables [2-4. When considering the same population and estimand, the only difference between tables that use BCa nonparametric bootstrap-based inference versus tables that use Wald-style inference is in the P(Reject *H*_0_) column.

**Table 2.** Results for the binary outcome and risk difference (RD) estimand in the hospitalized population. Wald-style inference is used for confidence intervals and hypothesis testing. “Effect” denotes the true estimand value; “MSE” denotes mean squared error; “Rel. Eff.” denotes relative efficiency which we approximate as the ratio of the MSE of the estimator under consideration to the MSE of the unadjusted estimator. In each block of four rows, the first two rows involve no treatment effect and the last two rows involve a benefit from treatment.

| $n$ | Estimator Type | Effect | P(reject $H_0$) | MSE | Bias | Variance | Rel. Eff. |
| --- | --- | --- | --- | --- | --- | --- | --- |
| 100 | Unadjusted | 0 | 0.043 | 0.010 | 0.003 | 0.010 | 1.000 |
| 100 | Adjusted | 0 | 0.056 | 0.009 | 0.004 | 0.009 | 0.844 |
| 100 | Unadjusted | -0.269 | 0.719 | 0.009 | 0.003 | 0.009 | 1.000 |
| 100 | Adjusted | -0.269 | 0.847 | 0.008 | 0.004 | 0.008 | 0.859 |
| 200 | Unadjusted | 0 | 0.031 | 0.005 | 0.003 | 0.005 | 1.000 |
| 200 | Adjusted | 0 | 0.041 | 0.004 | 0.004 | 0.004 | 0.885 |
| 200 | Unadjusted | -0.199 | 0.768 | 0.005 | 0.003 | 0.005 | 1.000 |
| 200 | Adjusted | -0.199 | 0.846 | 0.004 | 0.004 | 0.004 | 0.880 |
| 500 | Unadjusted | 0 | 0.047 | 0.002 | 0.001 | 0.002 | 1.000 |
| 500 | Adjusted | 0 | 0.051 | 0.002 | 0.000 | 0.002 | 0.878 |
| 500 | Unadjusted | -0.124 | 0.770 | 0.002 | 0.000 | 0.002 | 1.000 |
| 500 | Adjusted | -0.124 | 0.837 | 0.002 | 0.000 | 0.002 | 0.899 |
| 1000 | Unadjusted | 0 | 0.041 | 0.001 | 0.000 | 0.001 | 1.000 |
| 1000 | Adjusted | 0 | 0.042 | 0.001 | 0.000 | 0.001 | 0.860 |
| 1000 | Unadjusted | -0.090 | 0.796 | 0.001 | 0.000 | 0.001 | 1.000 |
| 1000 | Adjusted | -0.090 | 0.861 | 0.001 | 0.000 | 0.001 | 0.890 |

**Table 3.** Results for the binary outcome and risk difference (RD) estimand in the non-hospitalized population. BCa bootstrap is used for confidence intervals and hypothesis testing. “Effect” denotes the true estimand value; “MSE” denotes mean squared error; “Rel. Eff.” denotes relative efficiency which we approximate as the ratio of the MSE of the estimator under consideration to the MSE of the unadjusted estimator. In each block of four rows, the first two rows involve no treatment effect and the last two rows involve a benefit from treatment.

| $n$ | Estimator Type | Effect | P(reject $H_0$) | MSE | Bias | Variance | Rel. Eff. |
| --- | --- | --- | --- | --- | --- | --- | --- |
| 100 | Unadjusted | 0 | 0.029 | 0.008 | 0.003 | 0.008 | 1.000 |
| 100 | Adjusted | 0 | 0.060 | 0.008 | 0.001 | 0.008 | 0.918 |
| 100 | Unadjusted | -0.209 | 0.652 | 0.005 | 0.007 | 0.005 | 1.000 |
| 100 | Adjusted | -0.209 | 0.811 | 0.005 | 0.005 | 0.005 | 0.941 |
| 200 | Unadjusted | 0 | 0.043 | 0.004 | 0.000 | 0.004 | 1.000 |
| 200 | Adjusted | 0 | 0.059 | 0.004 | 0.001 | 0.004 | 0.885 |
| 200 | Unadjusted | -0.161 | 0.747 | 0.003 | -0.002 | 0.003 | 1.000 |
| 200 | Adjusted | -0.161 | 0.840 | 0.003 | -0.001 | 0.003 | 0.883 |
| 500 | Unadjusted | 0 | 0.042 | 0.002 | 0.000 | 0.002 | 1.000 |
| 500 | Adjusted | 0 | 0.057 | 0.002 | 0.000 | 0.002 | 0.887 |
| 500 | Unadjusted | -0.112 | 0.811 | 0.001 | 0.000 | 0.001 | 1.000 |
| 500 | Adjusted | -0.112 | 0.887 | 0.001 | 0.000 | 0.001 | 0.879 |
| 1000 | Unadjusted | 0 | 0.047 | 0.001 | 0.000 | 0.001 | 1.000 |
| 1000 | Adjusted | 0 | 0.056 | 0.001 | 0.000 | 0.001 | 0.920 |
| 1000 | Unadjusted | -0.073 | 0.712 | 0.001 | 0.001 | 0.001 | 1.000 |
| 1000 | Adjusted | -0.073 | 0.791 | 0.001 | 0.001 | 0.001 | 0.930 |

**Table 4.**
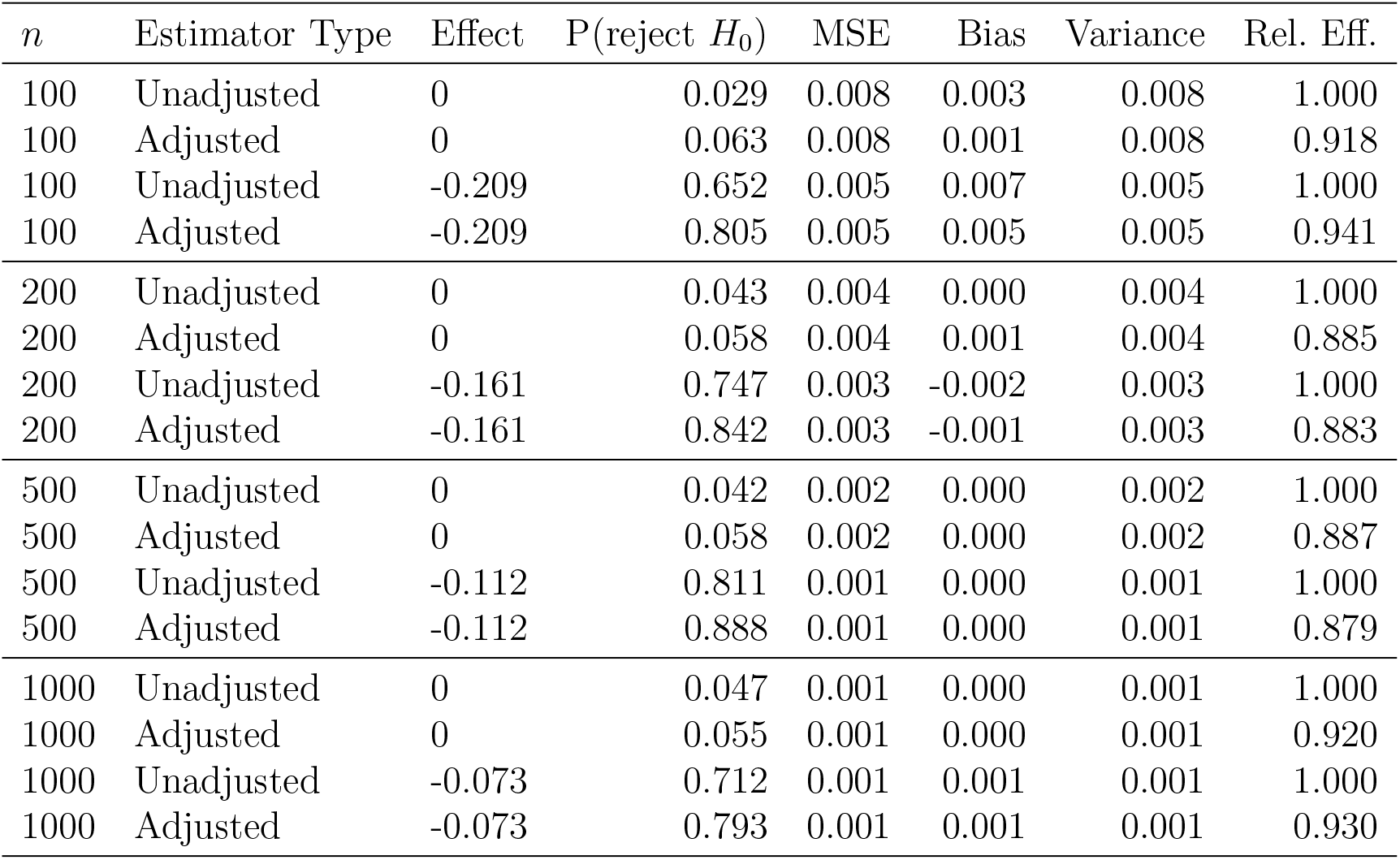
Results for the binary outcome and risk difference (RD) estimand in the non-hospitalized population. Wald-style inference is used for confidence intervals and hypothesis testing. “Effect” denotes the true estimand value; “MSE” denotes mean squared error; “Rel. Eff.” denotes relative efficiency which we approximate as the ratio of the MSE of the estimator under consideration to the MSE of the unadjusted estimator. In each block of four rows, the first two rows involve no treatment effect and the last two rows involve a benefit from treatment.

| $n$ | Estimator Type | Effect | P(reject $H_0$) | MSE | Bias | Variance | Rel. Eff. |
| --- | --- | --- | --- | --- | --- | --- | --- |
| 100 | Unadjusted | 0 | 0.029 | 0.008 | 0.003 | 0.008 | 1.000 |
| 100 | Adjusted | 0 | 0.063 | 0.008 | 0.001 | 0.008 | 0.918 |
| 100 | Unadjusted | -0.209 | 0.652 | 0.005 | 0.007 | 0.005 | 1.000 |
| 100 | Adjusted | -0.209 | 0.805 | 0.005 | 0.005 | 0.005 | 0.941 |
| 200 | Unadjusted | 0 | 0.043 | 0.004 | 0.000 | 0.004 | 1.000 |
| 200 | Adjusted | 0 | 0.058 | 0.004 | 0.001 | 0.004 | 0.885 |
| 200 | Unadjusted | -0.161 | 0.747 | 0.003 | -0.002 | 0.003 | 1.000 |
| 200 | Adjusted | -0.161 | 0.842 | 0.003 | -0.001 | 0.003 | 0.883 |
| 500 | Unadjusted | 0 | 0.042 | 0.002 | 0.000 | 0.002 | 1.000 |
| 500 | Adjusted | 0 | 0.058 | 0.002 | 0.000 | 0.002 | 0.887 |
| 500 | Unadjusted | -0.112 | 0.811 | 0.001 | 0.000 | 0.001 | 1.000 |
| 500 | Adjusted | -0.112 | 0.888 | 0.001 | 0.000 | 0.001 | 0.879 |
| 1000 | Unadjusted | 0 | 0.047 | 0.001 | 0.000 | 0.001 | 1.000 |
| 1000 | Adjusted | 0 | 0.055 | 0.001 | 0.000 | 0.001 | 0.920 |
| 1000 | Unadjusted | -0.073 | 0.712 | 0.001 | 0.001 | 0.001 | 1.000 |
| 1000 | Adjusted | -0.073 | 0.793 | 0.001 | 0.001 | 0.001 | 0.930 |

### C.3 Additional simulation studies for ordinal outcomes

We first present simulation results when using Wald-style inference for the population of hospitalized patients, in Tables 5-7 for the three ordinal estimands. Results were largely similar to those that used BCa nonparametric bootstrap-based inference as presented in the main text.

**Table 5.** Results for the ordinal outcome and difference in means (DIM) estimand in the hospitalized population. Wald-style inference is used for confidence intervals and hypothesis testing. “Effect” denotes the true estimand value; “MSE” denotes mean squared error; “Rel. Eff.” denotes relative efficiency which we approximate as the ratio of the MSE of the estimator under consideration to the MSE of the unadjusted estimator. In each block of four rows, the first two rows involve no treatment effect and the last two rows involve a benefit from treatment.

| $n$ | Estimator Type | Effect | P(reject $H_0$) | MSE | Bias | Variance | Rel. Eff. |
| --- | --- | --- | --- | --- | --- | --- | --- |
| 100 | Unadjusted | 0 | 0.067 | 0.023 | -0.005 | 0.023 | 1.000 |
| 100 | Adjusted | 0 | 0.065 | 0.019 | -0.007 | 0.019 | 0.822 |
| 100 | Unadjusted | 0.303 | 0.503 | 0.022 | -0.007 | 0.022 | 1.000 |
| 100 | Adjusted | 0.303 | 0.592 | 0.019 | -0.004 | 0.019 | 0.845 |
| 200 | Unadjusted | 0 | 0.042 | 0.010 | -0.002 | 0.010 | 1.000 |
| 200 | Adjusted | 0 | 0.047 | 0.009 | -0.003 | 0.009 | 0.862 |
| 200 | Unadjusted | 0.303 | 0.792 | 0.012 | -0.003 | 0.012 | 1.000 |
| 200 | Adjusted | 0.303 | 0.858 | 0.010 | 0.000 | 0.010 | 0.872 |
| 500 | Unadjusted | 0 | 0.060 | 0.005 | -0.001 | 0.005 | 1.000 |
| 500 | Adjusted | 0 | 0.057 | 0.004 | 0.000 | 0.004 | 0.837 |
| 500 | Unadjusted | 0.195 | 0.816 | 0.005 | 0.000 | 0.005 | 1.000 |
| 500 | Adjusted | 0.195 | 0.869 | 0.004 | 0.001 | 0.004 | 0.891 |
| 1000 | Unadjusted | 0 | 0.045 | 0.002 | 0.000 | 0.002 | 1.000 |
| 1000 | Adjusted | 0 | 0.044 | 0.002 | 0.000 | 0.002 | 0.849 |
| 1000 | Unadjusted | 0.136 | 0.826 | 0.002 | 0.000 | 0.002 | 1.000 |
| 1000 | Adjusted | 0.136 | 0.885 | 0.002 | 0.000 | 0.002 | 0.889 |

**Table 6.** Results for ordinal outcome and Mann Whitney (MW) estimand in the hospitalized population. Wald-style inference is used for confidence intervals and hypothesis testing. “Effect” denotes the true estimand value; “MSE” denotes mean squared error; “Rel. Eff.” denotes relative efficiency which we approximate as the ratio of the MSE of the estimator under consideration to the MSE of the unadjusted estimator. In each block of four rows, the first two rows involve no treatment effect and the last two rows involve a benefit from treatment.

| $n$ | Estimator Type | Effect | P(reject $H_0$) | MSE | Bias | Variance | Rel. Eff. |
| --- | --- | --- | --- | --- | --- | --- | --- |
| 100 | Unadjusted | 0.500 | 0.071 | 0.003 | -0.002 | 0.003 | 1.000 |
| 100 | Adjusted | 0.500 | 0.062 | 0.002 | -0.003 | 0.002 | 0.822 |
| 100 | Unadjusted | 0.627 | 0.607 | 0.002 | -0.002 | 0.002 | 1.000 |
| 100 | Adjusted | 0.627 | 0.696 | 0.002 | -0.002 | 0.002 | 0.852 |
| 200 | Unadjusted | 0.500 | 0.048 | 0.001 | -0.001 | 0.001 | 1.000 |
| 200 | Adjusted | 0.500 | 0.047 | 0.001 | -0.001 | 0.001 | 0.864 |
| 200 | Unadjusted | 0.627 | 0.917 | 0.001 | -0.001 | 0.001 | 1.000 |
| 200 | Adjusted | 0.627 | 0.959 | 0.001 | 0.000 | 0.001 | 0.878 |
| 500 | Unadjusted | 0.500 | 0.060 | 0.001 | 0.000 | 0.001 | 1.000 |
| 500 | Adjusted | 0.500 | 0.054 | 0.000 | 0.000 | 0.000 | 0.843 |
| 500 | Unadjusted | 0.582 | 0.926 | 0.001 | 0.000 | 0.001 | 1.000 |
| 500 | Adjusted | 0.582 | 0.950 | 0.000 | 0.000 | 0.000 | 0.905 |
| 1000 | Unadjusted | 0.500 | 0.044 | 0.000 | 0.000 | 0.000 | 1.000 |
| 1000 | Adjusted | 0.500 | 0.047 | 0.000 | 0.000 | 0.000 | 0.844 |
| 1000 | Unadjusted | 0.557 | 0.915 | 0.000 | 0.000 | 0.000 | 1.000 |
| 1000 | Adjusted | 0.557 | 0.940 | 0.000 | 0.000 | 0.000 | 0.890 |

**Table 7.** Results for the ordinal outcome and log-odds ratio (LOR) estimand in the hospitalized population. Wald-style inference is used for confidence intervals and hypothesis testing. “Effect” denotes the true estimand value; “MSE” denotes mean squared error; “Rel. Eff.” denotes relative efficiency which we approximate as the ratio of the MSE of the estimator under consideration to the MSE of the unadjusted estimator. In each block of four rows, the first two rows involve no treatment effect and the last two rows involve a benefit from treatment.

| $n$ | Estimator Type | Effect | P(reject $H_0$) | MSE | Bias | Variance | Rel. Eff. |
| --- | --- | --- | --- | --- | --- | --- | --- |
| 100 | Unadjusted | 0 | 0.053 | 0.185 | 0.018 | 0.185 | 1.000 |
| 100 | Adjusted | 0 | 0.057 | 0.153 | 0.021 | 0.152 | 0.824 |
| 100 | Unadjusted | -0.686 | 0.306 | 0.231 | 0.006 | 0.231 | 1.000 |
| 100 | Adjusted | -0.686 | 0.372 | 0.196 | 0.001 | 0.196 | 0.848 |
| 200 | Unadjusted | 0 | 0.042 | 0.080 | 0.004 | 0.081 | 1.000 |
| 200 | Adjusted | 0 | 0.044 | 0.069 | 0.007 | 0.069 | 0.854 |
| 200 | Unadjusted | -0.686 | 0.562 | 0.111 | 0.000 | 0.111 | 1.000 |
| 200 | Adjusted | -0.686 | 0.633 | 0.096 | -0.003 | 0.096 | 0.863 |
| 500 | Unadjusted | 0 | 0.060 | 0.035 | 0.002 | 0.035 | 1.000 |
| 500 | Adjusted | 0 | 0.065 | 0.029 | 0.000 | 0.029 | 0.826 |
| 500 | Unadjusted | -0.408 | 0.574 | 0.038 | -0.001 | 0.038 | 1.000 |
| 500 | Adjusted | -0.408 | 0.640 | 0.033 | -0.002 | 0.033 | 0.869 |
| 1000 | Unadjusted | 0 | 0.041 | 0.015 | 0.000 | 0.015 | 1.000 |
| 1000 | Adjusted | 0 | 0.047 | 0.013 | 0.000 | 0.013 | 0.851 |
| 1000 | Unadjusted | -0.278 | 0.577 | 0.016 | 0.000 | 0.016 | 1.000 |
| 1000 | Adjusted | -0.278 | 0.641 | 0.014 | 0.002 | 0.014 | 0.878 |

We also noted considerable numerical instabilities in implementations of the proportional odds model included in the MASS package (function polr), which led to our using the more stable implementation in the ordinal package (function clm) throughout. The latter function is the default in the drord package.

Tables 8-13 present simulation results for ordinal outcomes for the non-hospitalized population described in Section C.1 of the Supplementary Materials.

**Table 8.** Results for the ordinal outcome and difference in means (DIM) estimand in the non-hospitalized population. BCa bootstrap is used for confidence intervals and hypothesis testing. “Effect” denotes the true estimand value; “MSE” denotes mean squared error; “Rel. Eff.” denotes relative efficiency which we approximate as the ratio of the MSE of the estimator under consideration to the MSE of the unadjusted estimator. In each block of four rows, the first two rows involve no treatment effect and the last two rows involve a benefit from treatment.

| $n$ | Estimator Type | Effect | P(reject $H_0$) | MSE | Bias | Variance | Rel. Eff. |
| --- | --- | --- | --- | --- | --- | --- | --- |
| 100 | Unadjusted | 0 | 0.057 | 0.011 | -0.002 | 0.011 | 1.000 |
| 100 | Adjusted | 0 | 0.061 | 0.010 | -0.001 | 0.010 | 0.947 |
| 100 | Unadjusted | 0.193 | 0.504 | 0.009 | -0.006 | 0.009 | 1.000 |
| 100 | Adjusted | 0.193 | 0.535 | 0.008 | -0.004 | 0.008 | 0.950 |
| 200 | Unadjusted | 0 | 0.062 | 0.006 | 0.000 | 0.006 | 1.000 |
| 200 | Adjusted | 0 | 0.063 | 0.005 | -0.001 | 0.005 | 0.892 |
| 200 | Unadjusted | 0.193 | 0.816 | 0.005 | 0.003 | 0.005 | 1.000 |
| 200 | Adjusted | 0.193 | 0.844 | 0.004 | 0.003 | 0.004 | 0.915 |
| 500 | Unadjusted | 0 | 0.055 | 0.002 | 0.000 | 0.002 | 1.000 |
| 500 | Adjusted | 0 | 0.048 | 0.002 | 0.000 | 0.002 | 0.896 |
| 500 | Unadjusted | 0.125 | 0.791 | 0.002 | 0.000 | 0.002 | 1.000 |
| 500 | Adjusted | 0.125 | 0.838 | 0.002 | 0.000 | 0.002 | 0.894 |
| 1000 | Unadjusted | 0 | 0.054 | 0.001 | 0.000 | 0.001 | 1.000 |
| 1000 | Adjusted | 0 | 0.060 | 0.001 | 0.000 | 0.001 | 0.924 |
| 1000 | Unadjusted | 0.092 | 0.806 | 0.001 | -0.001 | 0.001 | 1.000 |
| 1000 | Adjusted | 0.092 | 0.832 | 0.001 | 0.000 | 0.001 | 0.948 |

**Table 9.** Results for the ordinal outcome and difference in means (DIM) estimand in the non-hospitalized population. Wald-style inference is used for confidence intervals and hypothesis testing. “Effect” denotes the true estimand value; “MSE” denotes mean squared error; “Rel. Eff.” denotes relative efficiency which we approximate as the ratio of the MSE of the estimator under consideration to the MSE of the unadjusted estimator. In each block of four rows, the first two rows involve no treatment effect and the last two rows involve a benefit from treatment.

| $n$ | Estimator Type | Effect | P(reject $H_0$) | MSE | Bias | Variance | Rel. Eff. |
| --- | --- | --- | --- | --- | --- | --- | --- |
| 100 | Unadjusted | 0 | 0.053 | 0.011 | -0.002 | 0.011 | 1.000 |
| 100 | Adjusted | 0 | 0.058 | 0.010 | -0.001 | 0.010 | 0.947 |
| 100 | Unadjusted | 0.193 | 0.495 | 0.009 | -0.006 | 0.009 | 1.000 |
| 100 | Adjusted | 0.193 | 0.562 | 0.008 | -0.004 | 0.008 | 0.950 |
| 200 | Unadjusted | 0 | 0.062 | 0.006 | 0.000 | 0.006 | 1.000 |
| 200 | Adjusted | 0 | 0.065 | 0.005 | -0.001 | 0.005 | 0.892 |
| 200 | Unadjusted | 0.193 | 0.831 | 0.005 | 0.003 | 0.005 | 1.000 |
| 200 | Adjusted | 0.193 | 0.863 | 0.004 | 0.003 | 0.004 | 0.915 |
| 500 | Unadjusted | 0 | 0.050 | 0.002 | 0.000 | 0.002 | 1.000 |
| 500 | Adjusted | 0 | 0.054 | 0.002 | 0.000 | 0.002 | 0.896 |
| 500 | Unadjusted | 0.125 | 0.790 | 0.002 | 0.000 | 0.002 | 1.000 |
| 500 | Adjusted | 0.125 | 0.846 | 0.002 | 0.000 | 0.002 | 0.894 |
| 1000 | Unadjusted | 0 | 0.052 | 0.001 | 0.000 | 0.001 | 1.000 |
| 1000 | Adjusted | 0 | 0.061 | 0.001 | 0.000 | 0.001 | 0.924 |
| 1000 | Unadjusted | 0.092 | 0.808 | 0.001 | -0.001 | 0.001 | 1.000 |
| 1000 | Adjusted | 0.092 | 0.845 | 0.001 | 0.000 | 0.001 | 0.948 |

**Table 10.** Results for ordinal outcome and Mann Whitney (MW) estimand in the non-hospitalized population. BCa bootstrap is used for confidence intervals and hypothesis testing. “Effect” denotes the true estimand value; “MSE” denotes mean squared error; “Rel. Eff.” denotes relative efficiency which we approximate as the ratio of the MSE of the estimator under consideration to the MSE of the unadjusted estimator. In each block of four rows, the first two rows involve no treatment effect and the last two rows involve a benefit from treatment.

| $n$ | Estimator Type | Effect | P(reject $H_0$) | MSE | Bias | Variance | Rel. Eff. |
| --- | --- | --- | --- | --- | --- | --- | --- |
| 100 | Unadjusted | 0.500 | 0.051 | 0.002 | -0.001 | 0.002 | 1.000 |
| 100 | Adjusted | 0.500 | 0.050 | 0.002 | -0.001 | 0.002 | 0.939 |
| 100 | Unadjusted | 0.594 | 0.642 | 0.001 | -0.003 | 0.001 | 1.000 |
| 100 | Adjusted | 0.594 | 0.682 | 0.001 | -0.002 | 0.001 | 0.945 |
| 200 | Unadjusted | 0.500 | 0.062 | 0.001 | 0.000 | 0.001 | 1.000 |
| 200 | Adjusted | 0.500 | 0.058 | 0.001 | -0.001 | 0.001 | 0.893 |
| 200 | Unadjusted | 0.594 | 0.930 | 0.001 | 0.001 | 0.001 | 1.000 |
| 200 | Adjusted | 0.594 | 0.943 | 0.001 | 0.001 | 0.001 | 0.915 |
| 500 | Unadjusted | 0.500 | 0.052 | 0.000 | 0.000 | 0.000 | 1.000 |
| 500 | Adjusted | 0.500 | 0.051 | 0.000 | 0.000 | 0.000 | 0.898 |
| 500 | Unadjusted | 0.561 | 0.883 | 0.000 | 0.000 | 0.000 | 1.000 |
| 500 | Adjusted | 0.561 | 0.922 | 0.000 | 0.000 | 0.000 | 0.893 |
| 1000 | Unadjusted | 0.500 | 0.055 | 0.000 | 0.000 | 0.000 | 1.000 |
| 1000 | Adjusted | 0.500 | 0.062 | 0.000 | 0.000 | 0.000 | 0.929 |
| 1000 | Unadjusted | 0.544 | 0.898 | 0.000 | 0.000 | 0.000 | 1.000 |
| 1000 | Adjusted | 0.544 | 0.919 | 0.000 | 0.000 | 0.000 | 0.954 |

**Table 11.** Results for ordinal outcome and Mann Whitney (MW) estimand in the non-hospitalized population. Wald-style inference is used for confidence intervals and hypothesis testing. “Effect” denotes the true estimand value; “MSE” denotes mean squared error; “Rel. Eff.” denotes relative efficiency which we approximate as the ratio of the MSE of the estimator under consideration to the MSE of the unadjusted estimator. In each block of four rows, the first two rows involve no treatment effect and the last two rows involve a benefit from treatment.

| $n$ | Estimator Type | Effect | P(reject $H_0$) | MSE | Bias | Variance | Rel. Eff. |
| --- | --- | --- | --- | --- | --- | --- | --- |
| 100 | Unadjusted | 0.500 | 0.057 | 0.002 | -0.001 | 0.002 | 1.000 |
| 100 | Adjusted | 0.500 | 0.069 | 0.002 | -0.001 | 0.002 | 0.939 |
| 100 | Unadjusted | 0.594 | 0.538 | 0.001 | -0.003 | 0.001 | 1.000 |
| 100 | Adjusted | 0.594 | 0.612 | 0.001 | -0.002 | 0.001 | 0.945 |
| 200 | Unadjusted | 0.500 | 0.068 | 0.001 | 0.000 | 0.001 | 1.000 |
| 200 | Adjusted | 0.500 | 0.069 | 0.001 | -0.001 | 0.001 | 0.893 |
| 200 | Unadjusted | 0.594 | 0.901 | 0.001 | 0.001 | 0.001 | 1.000 |
| 200 | Adjusted | 0.594 | 0.924 | 0.001 | 0.001 | 0.001 | 0.915 |
| 500 | Unadjusted | 0.500 | 0.050 | 0.000 | 0.000 | 0.000 | 1.000 |
| 500 | Adjusted | 0.500 | 0.057 | 0.000 | 0.000 | 0.000 | 0.898 |
| 500 | Unadjusted | 0.561 | 0.861 | 0.000 | 0.000 | 0.000 | 1.000 |
| 500 | Adjusted | 0.561 | 0.910 | 0.000 | 0.000 | 0.000 | 0.893 |
| 1000 | Unadjusted | 0.500 | 0.054 | 0.000 | 0.000 | 0.000 | 1.000 |
| 1000 | Adjusted | 0.500 | 0.060 | 0.000 | 0.000 | 0.000 | 0.929 |
| 1000 | Unadjusted | 0.544 | 0.885 | 0.000 | 0.000 | 0.000 | 1.000 |
| 1000 | Adjusted | 0.544 | 0.911 | 0.000 | 0.000 | 0.000 | 0.954 |

**Table 12.**
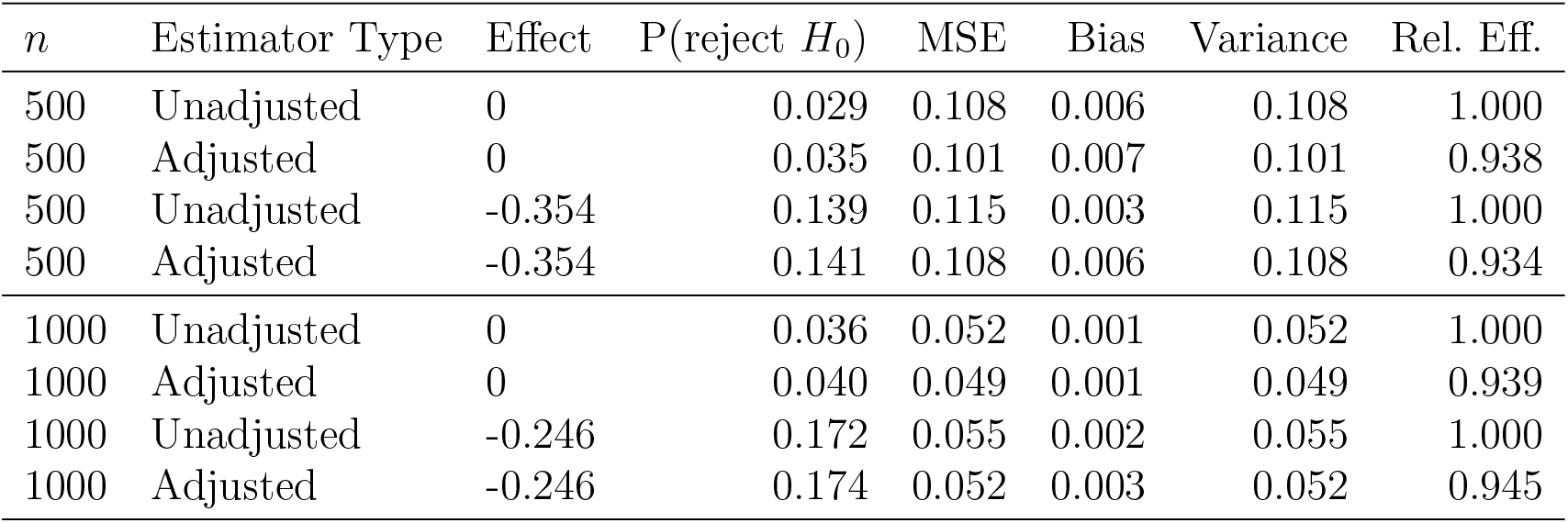
Results for the ordinal outcome and log-odds ratio (LOR) estimand in the non-hospitalized population. BCa bootstrap is used for confidence intervals and hypothesis testing. “Effect” denotes the true estimand value; “MSE” denotes mean squared error; “Rel. Eff.” denotes relative efficiency which we approximate as the ratio of the MSE of the estimator under consideration to the MSE of the unadjusted estimator. In each block of four rows, the first two rows involve no treatment effect and the last two rows involve a benefit from treatment.

**Table 13.** Results for the ordinal outcome and log-odds ratio (LOR) estimand in the non-hospitalized population. Wald-style inference is used for confidence intervals and hypothesis testing. “Effect” denotes the true estimand value; “MSE” denotes mean squared error; “Rel. Eff.” denotes relative efficiency which we approximate as the ratio of the MSE of the estimator under consideration to the MSE of the unadjusted estimator. In each block of four rows, the first two rows involve no treatment effect and the last two rows involve a benefit from treatment.

| $n$ | Estimator Type | Effect | P(reject $H_0$) | MSE | Bias | Variance | Rel. Eff. |
| --- | --- | --- | --- | --- | --- | --- | --- |
| 500 | Unadjusted | 0 | 0.031 | 0.108 | 0.006 | 0.108 | 1.000 |
| 500 | Adjusted | 0 | 0.040 | 0.101 | 0.007 | 0.101 | 0.938 |
| 500 | Unadjusted | -0.354 | 0.184 | 0.115 | 0.003 | 0.115 | 1.000 |
| 500 | Adjusted | -0.354 | 0.208 | 0.108 | 0.006 | 0.108 | 0.934 |
| 1000 | Unadjusted | 0 | 0.045 | 0.052 | 0.001 | 0.052 | 1.000 |
| 1000 | Adjusted | 0 | 0.063 | 0.049 | 0.001 | 0.049 | 0.939 |
| 1000 | Unadjusted | -0.246 | 0.180 | 0.055 | 0.002 | 0.055 | 1.000 |
| 1000 | Adjusted | -0.246 | 0.210 | 0.052 | 0.003 | 0.052 | 0.945 |

### C.4 Additional simulation studies for time-to-event outcomes

We present results for the difference in restricted mean survival times (RMST) at 14 days estimand in the hospitalized population, when the adjusted estimator uses only age and sex (Table 14). Results are also presented for the difference of survival probabilities (RD) at 7 days estimand in the hospitalized population (when the adjusted estimator uses all six baseline variables from Section 4.2.3) in Table 15

**Table 14.** Results for difference in restricted mean survival times (RMST) at 14 days estimand in hospitalized population, when the adjusted estimator uses only age and sex. Confidence intervals and hypothesis tests are Wald-style. “Effect” denotes the true estimand value; “MSE” denotes mean squared error; “Rel. Eff.” denotes relative efficiency which we approximate as the ratio of the MSE of the estimator under consideration to the MSE of the unadjusted estimator. In each block of four rows, the first two rows involve no treatment effect and the last two rows involve a benefit from treatment.

| $n$ | Estimator Type | Effect | P(reject $H_0$) | MSE | Bias | Variance | Rel. Eff. |
| --- | --- | --- | --- | --- | --- | --- | --- |
| 100 | Unadjusted | 0 | 0.011 | 0.781 | 0.018 | 0.780 | 1.000 |
| 100 | Adjusted | 0 | 0.015 | 0.771 | 0.014 | 0.771 | 0.987 |
| 100 | Unadjusted | 1.06 | 0.085 | 0.570 | -0.265 | 0.500 | 1.000 |
| 100 | Adjusted | 1.06 | 0.098 | 0.570 | -0.263 | 0.501 | 1.000 |
| 200 | Unadjusted | 0 | 0.048 | 0.481 | -0.013 | 0.481 | 1.000 |
| 200 | Adjusted | 0 | 0.050 | 0.476 | -0.014 | 0.476 | 0.989 |
| 200 | Unadjusted | 1.06 | 0.326 | 0.328 | -0.145 | 0.307 | 1.000 |
| 200 | Adjusted | 1.06 | 0.337 | 0.326 | -0.141 | 0.306 | 0.995 |
| 500 | Unadjusted | 0 | 0.050 | 0.201 | -0.003 | 0.201 | 1.000 |
| 500 | Adjusted | 0 | 0.049 | 0.196 | -0.003 | 0.196 | 0.975 |
| 500 | Unadjusted | 1.06 | 0.729 | 0.151 | -0.070 | 0.146 | 1.000 |
| 500 | Adjusted | 1.06 | 0.742 | 0.147 | -0.069 | 0.143 | 0.978 |
| 1000 | Unadjusted | 0 | 0.048 | 0.100 | 0.001 | 0.100 | 1.000 |
| 1000 | Adjusted | 0 | 0.047 | 0.096 | 0.001 | 0.096 | 0.963 |
| 1000 | Unadjusted | 1.06 | 0.959 | 0.079 | -0.060 | 0.076 | 1.000 |
| 1000 | Adjusted | 1.06 | 0.963 | 0.077 | -0.060 | 0.073 | 0.972 |

**Table 15.** Results for difference of survival probabilities (RD) at 7 days estimand in hospitalized population when the adjusted estimator uses all six baseline variables from Section 4-2.3. Confidence intervals and hypothesis tests are Wald-style. “Effect” denotes the true estimand value; “MSE” denotes mean squared error; “Rel. Eff.” denotes relative efficiency which we approximate as the ratio of the MSE of the estimator under consideration to the MSE of the unadjusted estimator. In each block of four rows, the first two rows involve no treatment effect and the last two rows involve a benefit from treatment.

| Sample Size | Estimator Type | Effect | P(reject $H_0$) | MSE | Bias | Variance | Rel. Eff. |
| --- | --- | --- | --- | --- | --- | --- | --- |
| 100 | Unadjusted | 0 | 0.052 | 0.007 | 0.001 | 0.008 | 1.000 |
| 100 | Adjusted | 0 | 0.065 | 0.007 | 0.001 | 0.007 | 0.935 |
| 100 | Unadjusted | 0.087 | 0.185 | 0.007 | -0.002 | 0.007 | 1.000 |
| 100 | Adjusted | 0.087 | 0.209 | 0.006 | -0.001 | 0.006 | 0.973 |
| 200 | Unadjusted | 0 | 0.050 | 0.004 | -0.001 | 0.004 | 1.000 |
| 200 | Adjusted | 0 | 0.058 | 0.003 | -0.001 | 0.003 | 0.869 |
| 200 | Unadjusted | 0.087 | 0.316 | 0.003 | -0.003 | 0.003 | 1.000 |
| 200 | Adjusted | 0.087 | 0.357 | 0.003 | -0.003 | 0.003 | 0.904 |
| 500 | Unadjusted | 0 | 0.053 | 0.002 | 0.001 | 0.002 | 1.000 |
| 500 | Adjusted | 0 | 0.052 | 0.001 | 0.001 | 0.001 | 0.838 |
| 500 | Unadjusted | 0.087 | 0.648 | 0.001 | -0.002 | 0.001 | 1.000 |
| 500 | Adjusted | 0.087 | 0.717 | 0.001 | -0.002 | 0.001 | 0.858 |
| 1000 | Unadjusted | 0 | 0.052 | 0.001 | 0.001 | 0.001 | 1.000 |
| 1000 | Adjusted | 0 | 0.051 | 0.001 | 0.001 | 0.001 | 0.833 |
| 1000 | Unadjusted | 0.087 | 0.918 | 0.001 | -0.002 | 0.001 | 1.000 |
| 1000 | Adjusted | 0.087 | 0.947 | 0.001 | -0.002 | 0.001 | 0.851 |

## D. Code availability

### D.1 Simulation code

All code needed to reproduce the simulations for ordinal and binary data is available on GitHub (https://github.com/mrosenblum/COVID-19-RCT-STAT-TOOLS). The code for the survival simulations is also included in that repository. However, because the simulation is based on private data from Weill Cornell Medicine, the results of the simulation reported in the manuscript are not reproducible based on the available code. We provide a simulated dataset (not based on real data) with the same structure of the real dataset. This dataset can be used to run the simulation code.

### D.2 R packages

The drord package (available at https://github.com/benkeser/drord) implements the proposed estimators for ordinal outcomes and can also be used for analyzing binary outcomes. The package vignette(https://benkeser.github.io/drord/articles/using_drord.html) describes implementation of the estimators and all available options in the package. In particular, the package includes: bootstrap-based and closed-form inference for all estimands described here-in, as well as for the treatment-specific PMFs and CDFs; a fully nonparametric covariate-adjusted estimator that uses stratification to estimate the covariate-conditional PMF and estimators; and a plotting method for visualizing covariate-adjusted estimates of the treatment-specific PMFs and CDFs that includes pointwise confidence intervals and simultaneous confidence bands.

The survtmlerct package, available at https://github.com/idiazst/survtmlerct, implements the targeted minimum loss based estimator for the RMST of Díaz et al. (2019). The package also implements an analogous estimator for the risk difference RD, as well as unadjusted counterparts for both the RMST and the RD. Standard errors are computed using the influence function of the estimators, and Wald-type confidence intervals are implemented. The functions in the package can incorporate any user-provided, preliminary estimates of the outcome and hazard functions, including parametric and data-adaptive estimates that use model selection. The help command applied to the specific functions of the package gives examples of the estimators.

